# SickMix: Temporal Changes in Social Contact Patterns among People with Acute Infection and Their Close Contacts

**DOI:** 10.64898/2026.09.21.26362162

**Authors:** Jessica C. Ibiebele, Aarushi Tuli, Grissel Lopes, Gina Lombard, Anne Shapiro, Judy Donald, Mark A. Schmidt, Maria Litvinova, Dehao Chen, Samuel M. Jenness, Benjamin A. Lopman, Kayoko Shioda

## Abstract

Infected individuals drive infectious disease transmission, yet empirical data on social interactions during acute infection remain limited. We conducted a prospective longitudinal study of 1,000 medically attended acute gastroenteritis and acute respiratory infection cases and 709 household members in the northwestern United States from 2024 to 2025 to quantify illness-associated changes in social contacts according to age and social setting. Cases substantially reduced social contacts during peak illness; the weighted mean number of contacts on the index date was 6.7 (95% confidence interval (CI): 5.9–7.7), which increased to 13.1 (95% CI: 11.6–14.6) after one week and 15.3 (95% CI: 13.5–17.2) after two weeks. On average, the number of contacts increased 2.0-fold (95% CI: 1.8–2.2) over the two-week follow-up. The largest increase in social contacts over time was observed among school-aged children (5–17 years), driven primarily by increases in contacts with others in the same age group. These findings suggest that behavioral responses during acute infection could substantially influence transmission dynamics and intervention evaluation.

**Significance:** Mathematical models of infectious disease transmission guide public health decisions, from vaccination strategies to isolation policies. These models typically either assume that sick people interact with others as often as healthy people do or assume complete isolation during illness, neither of which likely reflects reality. By following 1,000 people with respiratory or gastrointestinal infections over two weeks, we found that they sharply reduced their social contacts during peak illness and doubled them over the subsequent two weeks as they recovered. Incorporating this behavior into transmission models could substantially change the predicted course of outbreaks and the estimated benefit of interventions. Our data enable models that capture how people behave when sick, supporting more reliable public health planning.

## Introduction

Human behavior plays a critical role in the transmission of infectious diseases (*1*). Social contact patterns determine how quickly and widely infections may spread among the population, and thus, are fundamental inputs in mathematical models for infectious diseases (*1*). However, most transmission models make extreme assumptions that may not reflect real-world behavior, such as assuming either fixed contact patterns over time or complete isolation during illness, largely because of the lack of empirical data on social contacts during illness (*2*, *3*). Since the early 2010s, infectious disease models have increasingly incorporated individual behavioral change, such as compliance with public health orders or voluntary preventive measures taken in response to information about disease threats (*4–6*). These models suggest that behavioral responses to perceived infection risk can substantially alter projected transmission dynamics and estimates of intervention effectiveness. However, most of these models focus on behavior that prevents infection rather than changes in behavior among acutely infected individuals, and they are largely theoretical, with limited empirical data on social contacts.

Although many studies have characterized social contact patterns, important gaps remain in understanding how social contacts change as individuals become ill and recover. For example, a landmark study collected comprehensive data on social contact patterns across age groups and settings in eight European countries in 2008 (*7*), which has informed many infectious disease models and policies (*8–16*). While these classic studies, among others, provide crucial baseline contact data, they generally do not capture within-person contact changes over the course of illness; moreover, they generally represent healthy individuals or those whose health status is unknown (*17*). More recently, during the COVID-19 pandemic, various surveys were conducted to understand how individuals changed their behaviors and social contact patterns, influenced by public health orders and community-level disease burden (*18–21*). These studies measured population-level behavior changes driven by the pandemic context and interventions, rather than behavior changes driven by an individual’s own illness. Thus, there remains a lack of empirical longitudinal data on how social contact patterns change within individuals throughout the course of acute illness and recovery.

Longitudinal data on social contact patterns have been more readily available for chronic infectious diseases, such as HIV and tuberculosis, than for acute infectious diseases (*22*, *23*). Transmission models for HIV and other sexually transmitted infections increasingly incorporate risk perception and associated behavior change (*24*, *25*). In contrast, only two studies have longitudinally assessed social contact patterns among individuals experiencing acute infection. A study in the United Kingdom found that individuals with acute respiratory infection (ARI) significantly reduced their social contacts during illness, reporting an average of 3.8 daily contacts on the sick day compared to 14.9 contacts at the two-week follow up when healthy (*26*). In contrast, another study conducted in Malawi observed relatively stable numbers of social contacts among cases over the two-week follow-up period (*27*). Differences between these two studies suggested that illness-associated changes in social contact patterns may vary across populations because of differences in demographic characteristics, social structure, cultural norms, or contextual factors. Moreover, both studies were conducted before the COVID-19 pandemic, highlighting the need for contemporary longitudinal data on social contact patterns during acute illness, particularly as perceptions of social distancing during illness may have changed in response to pandemic interventions. Finally, while the Malawi study also collected data from caregivers, longitudinal social contact data from close contacts exposed to acute infection remain scarce.

Therefore, we conducted the “SickMix” study, a prospective longitudinal study of individuals experiencing ARI or acute gastroenteritis (AGE) and their household members in the U.S. from 2024 to 2025. The primary objective was to quantify temporal changes in social contact patterns among individuals with ARI or AGE over two weeks and evaluate their implications for infectious disease transmission modeling. We also characterized temporal changes in social contacts among exposed household members of these cases. In addition, we assessed changes in infection mitigation behaviors among both cases and household members during acute infection and exposure. Finally, we created publicly available age-stratified, illness phase-specific contact matrices that can be incorporated into transmission models, potentially enabling more realistic epidemic simulations and evaluation of public health interventions.

## Materials and Methods

### SickMix study overview

The SickMix study was a prospective longitudinal study of medically attended ARI and AGE cases of all ages and their household members. The primary outcome was the mean number of social contacts on the day of peak illness (referred to as the index date), one week later, and two weeks later among ARI and AGE cases. Household members were also recruited and completed social contact surveys on the index date and one week later. Longitudinal social contact data were subsequently used in transmission modeling to understand the implications of social contact patterns over the course of illness for disease transmission and intervention evaluation.

### Study site

The SickMix study was conducted in collaboration with Kaiser Permanente Northwest (KPNW), an integrated healthcare delivery system serving approximately 619,000 members as of 2022. KPNW members comprised approximately 24% of the population in Northwest Oregon and Southwest Washington. The demographic characteristics of KPNW members were similar to those of the general population in the region (*28*).

### AGE and ARI case definition, recruitment, and eligibility criteria

A case was defined as an individual of any age who sought medical care within the KPNW system for AGE or ARI symptoms. Cases were identified through electronic health records using International Classification of Diseases, 10th Revision (ICD-10) codes or chief complaint codes associated with symptoms of ARI (coughing, sore throat, sneezing, runny nose, and/or difficulty breathing; **Supplementary Table 1**) or AGE (diarrhea and/or vomiting; **Supplementary Table 2**) within the previous 24 hours, on a daily basis. Recruitment of AGE and ARI cases occurred from March 18 to June 30, 2024, and from October 14, 2024, to March 31, 2025. Participant recruitment was paused over the summer due to the seasonal decline in AGE and ARI cases in the U.S.

Identified eligible AGE and ARI cases were automatically sent recruitment emails within 24 hours of their healthcare visit. To minimize recall bias, participants were excluded from the study if they did not respond within six days of receiving the recruitment email. AGE and ARI cases were asked to complete an online enrollment survey, which further determined their eligibility (**Supplementary Methods**).

### Surveys for AGE and ARI cases

Once AGE and ARI cases were deemed eligible, they provided socio-demographic information, living settings, education level, and total household income before taxes in the enrollment survey. For participants under 18 years of age, surveys could be completed by a parent or guardian. Age and sex were extracted from electronic health records. Respondents also reported whether they currently attend school and, if so, whether they attend classes in person or remotely. Participants aged ≥18 years were additionally asked about their employment status. If employed, respondents reported their primary work setting (outside the home, inside the home, or hybrid) and their occupation (health care, food service, education, or other).

Next, cases were asked to complete a longitudinal online social contact survey for three dates: (1) the day they felt most sick (index date), (2) one week later, and (3) two weeks later. If they felt equally sick on multiple days, they were instructed to select the most recent day as the index date. Social contacts were defined as any physical contact (e.g., handshake, hug), conversational contact (close-range verbal conversation), or close-proximity contact within six feet. Participants were asked to report contacts lasting more than five minutes to focus on substantive social interactions (*7*, *29*, *30*), reduce potential recall error from brief incidental encounters (*31*), and minimize response burden among acutely ill participants (**Supplementary Methods**).

The survey captured information on both household contacts at home and non-household contacts occurring at home, work, school, or general community. Participants reported the number of individuals they interacted with, categorized by age group (<5, 5–17, 18–29, 30–44, 45–64, and ≥65 years), duration of contact (5–9 minutes, 10–59 minutes, or ≥60 minutes), and setting (home, work, school, or general community).

The survey also collected data on infection mitigation behaviors on each survey day, such as washing hands and wearing masks more frequently, reducing the number of contacts, increasing physical distance, or missing work or school. The one-week and two-week follow-up surveys began with a self-reported health status question, in which cases indicated whether they still felt very sick, somewhat sick, a little sick, or had fully recovered.

The survey instrument is available in the **Supplementary Materials**.

### Household member recruitment and surveys

Household members were defined as people, regardless of familial relationship, who live in the same house, sharing the common area (e.g., kitchen and/or bathroom) (**Supplementary Methods**). Household members filled out the same social contact surveys used for AGE and ARI cases on the index date (i.e., the day the AGE or ARI case in their household felt most sick) and at the one-week follow-up. On each survey date, household members self-reported their own health status, indicating whether they felt very sick, somewhat sick, a little sick, or not sick. As with AGE and ARI cases, surveys could be completed by a parent or guardian for household members under 18 years of age.

### Contact matrices and survey weighting

Contact matrices were calculated by dividing the age group-specific number of contacts reported on each date by the number of participants in that age group. We excluded respondents who reported more than 20 contacts at home or more than 250 contacts at work, school, or other community settings because such large numbers were deemed implausible given that participants were only asked to report physical, conversational or close-proximity contacts lasting more than five minutes. This resulted in the exclusion of five ARI or AGE cases and one household member who reported more than 20 contacts with household members at home, in addition to four cases and three household members who reported more than 250 contacts with non-household members at a single non-household location. These thresholds were modified in sensitivity analyses to assess their impact on the results (see “Sensitivity Analyses”).

To make our study population representative of medically attended ARI and AGE cases in the U.S., we applied post-stratification weights based on the joint distribution of sex, age group, and disease type (ARI or AGE) in the 2023 IBM MarketScan Commercial Claims and Encounters Database and Medicare Supplemental Database. For each stratum defined by sex, age group, and disease type, the post-stratification weight was calculated as the ratio of the number of medically attended cases in the MarketScan/Medicare database to the corresponding number of participants enrolled in the SickMix study.

To account for uncertainty, we generated 500 bootstrap resamples by sampling cases with replacement, using the post-stratification weights as sampling probabilities. Summary statistics and statistical models were run separately for each bootstrap resample. We reported the mean of the resulting bootstrap estimates, with 2.5th and 97.5th percentiles of the bootstrap distribution as the 95% confidence interval. Unless otherwise specified, all results reported for AGE and ARI cases are bootstrap-weighted estimates.

### Statistical analysis

We fit mixed-effects negative binomial regression models to assess temporal changes in the number of social contacts from the index date to follow-up time points. The outcome variable was the number of social contacts reported each day. A random slope for participant was included to account for within-subject clustering. The primary predictor was time points (index date, one-week follow-up, and two-week follow-up).

### Sensitivity analyses

Our primary analyses focused on cases who completed all three longitudinal social contact surveys. As a sensitivity analysis, we calculated the mean number of contacts at each survey date among all participants who completed that specific survey, without restricting the analysis to those who completed subsequent follow-up surveys. We also assessed the impact of the threshold used to identify outliers in the reported number of contacts. In this sensitivity analysis, we applied a threshold of 50 contacts to the total number of reported contacts, truncated reported contacts exceeding this threshold to 50, and repeated the analysis.

All analyses were conducted using R version 4.5.2 (R Core Team, Vienna, Austria).

### Ethical review and participant incentives

The study protocol was approved by the Kaiser Permanente Interregional Institutional Review Board (IRB), as well as the IRBs of Emory University and Boston University (IRB# 2130008). Electronic informed consent was obtained from adult participants through an online form prior to survey participation. For participants under 18 years of age, a parent or guardian provided permission; for children aged 7 years or older, the parent or guardian was also asked to confirm that the child wished to participate. Cases and household members received a total of $20 and $15 gift cards, respectively, upon completion of the longitudinal surveys. All data were de-identified before analysis.

## Results

### Characteristics of AGE and ARI cases

A total of 1,401 cases completed the first social contact survey for the index date, 1,109 completed the one-week follow-up survey, and 1,009 cases completed the two-week follow-up survey. Nine cases were excluded from the analysis because their reported social contacts exceeded the threshold. Consequently, the main analysis included data from 1,000 AGE and ARI cases who completed all three longitudinal surveys. There were no significant differences in demographic characteristics between cases who provided consent and those who completed all three surveys (**Supplementary Table 3**).

In the weighted bootstrap resamples, cases were predominantly female (58.6%), aged 5–17 years (24.1%), White (76.2%), and non-Hispanic (82.1%) (**Supplementary Table 4**). Most cases lived in a house (80.8%). Of those enrolled in school, the majority attended in person (89.4%). Among individuals aged ≥18 years, 78.1% were employed at the time of the survey, with many reporting that they primarily worked outside their home. Regarding the type of healthcare encounter, 32.0% used telehealth, 66.0% sought ambulatory care, and 1.9% visited emergency departments.

### Temporal changes in social contact behaviors among AGE and ARI cases

Among 1,000 AGE and ARI cases who completed all longitudinal surveys, the weighted mean number of social contacts on the index date was 6.7 (95% CI: 5.9–7.7), which increased to 13.1 (95% CI: 11.6–14.6) at the one-week follow-up and 15.3 (95% CI: 13.5–17.2) at the two-week follow-up (**Figure 1**). On average, the number of contacts at the 2-week follow-up was 2.0 times (95% CI: 1.8–2.2) higher than on the index date. An increase was observed even among cases who self-reported that they remained sick at one and two-week follow ups (**Supplementary Figure 1**). The largest increase in social contacts over time was observed among school-aged children (5–17 years) (**Figure 2**), driven primarily by increases in contacts with others in the same age group (**Figure 3**). Regarding contact settings, the average number of contacts increased over the course of infection at school and daycare, while contacts at home and work remained stable (**Figure 4**). Compared to changes in the number of contacts, changes in the duration of contacts were subtle (**Figure 4**).

**Figure 1.**
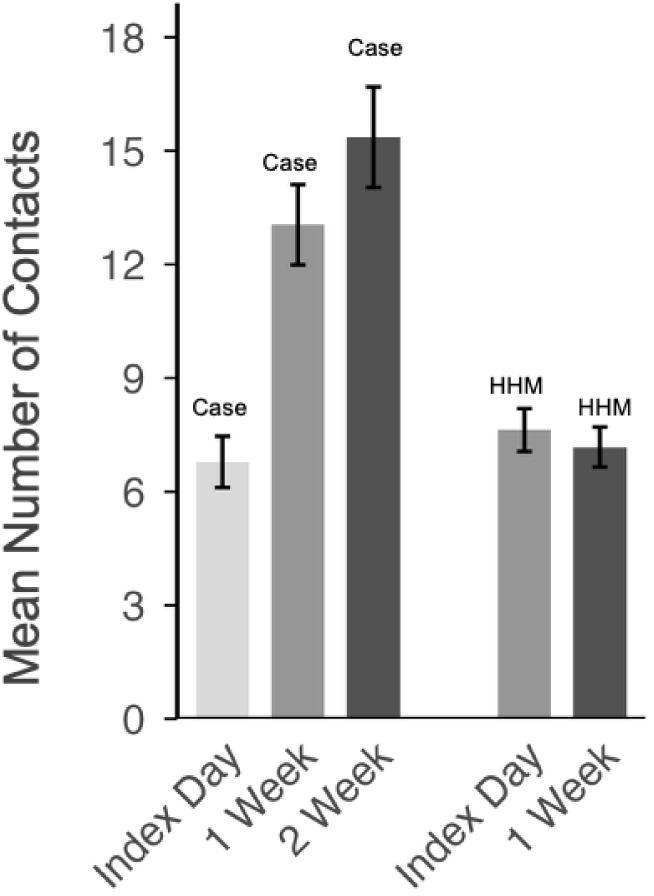
Mean number of social contacts reported by acute gastroenteritis and acute respiratory infection cases and their household members over the course of acute infection. Abbreviation: HHM: household members. The figure displays weighted mean contacts reported by cases from 500 bootstrap resampled iterations of N=1,000 (left columns) and unweighted mean contacts reported by household members (right columns). Vertical lines represent error bars—the standard deviation of the bootstrap means for cases and standard errors for household members. Data from study participants that completed all longitudinal surveys are included.

**Figure 2.**
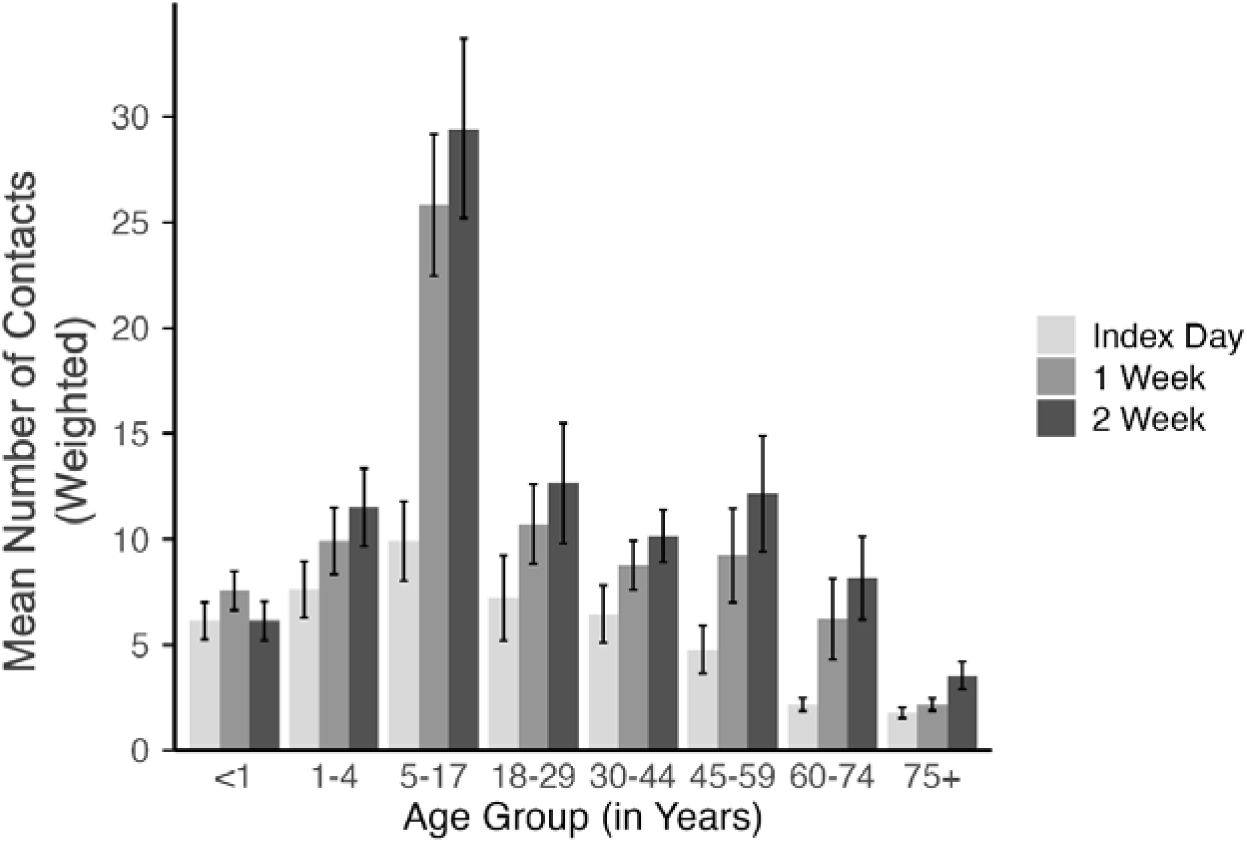
Weighted mean number of social contacts reported by acute gastroenteritis and acute respiratory infection cases over time by age group. This figure displays weighted mean contacts for cases from 500 bootstrap resampled iterations of N=1,000. Vertical lines represent the standard deviation of the bootstrap means. Data from study participants that completed all longitudinal surveys are included.

**Figure 3.**
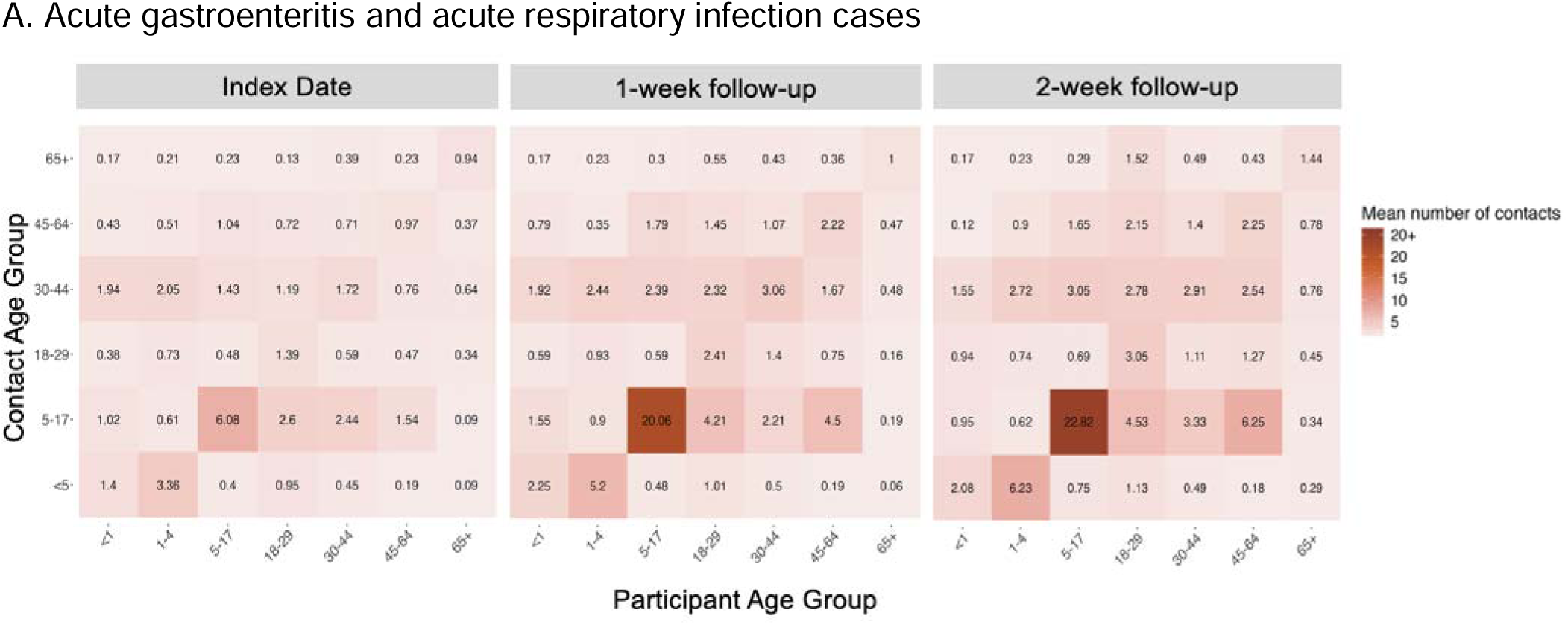
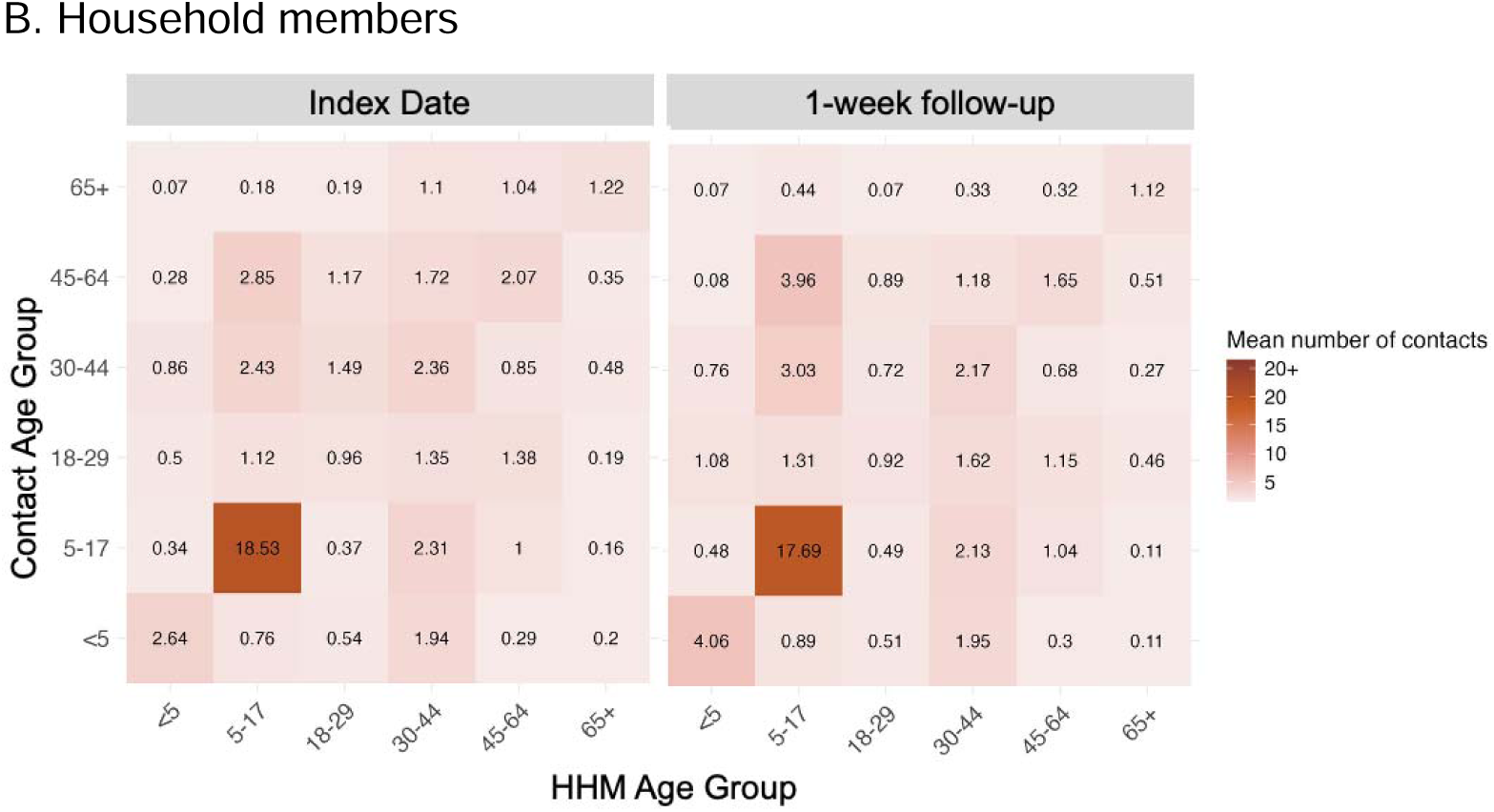
Age-stratified social contact matrices (mean number of contacts) reported by acute gastroenteritis and acute respiratory infection cases and their household members over the course of acute infection, with the greatest increase in contacts among school-aged children (5-17 years) with peers. A. Acute gastroenteritis and acute respiratory infection cases B. Household members Abbreviation: HHM: household members. Panel A displays weighted contact matrices (weighted mean number of contacts) for acute gastroenteritis and acute respiratory infection cases and their contacts from 500 bootstrap resampled iterations of N=1,000. Panel B displays contact matrices (mean number of contacts) for household contacts (N=709).

**Figure 4.**
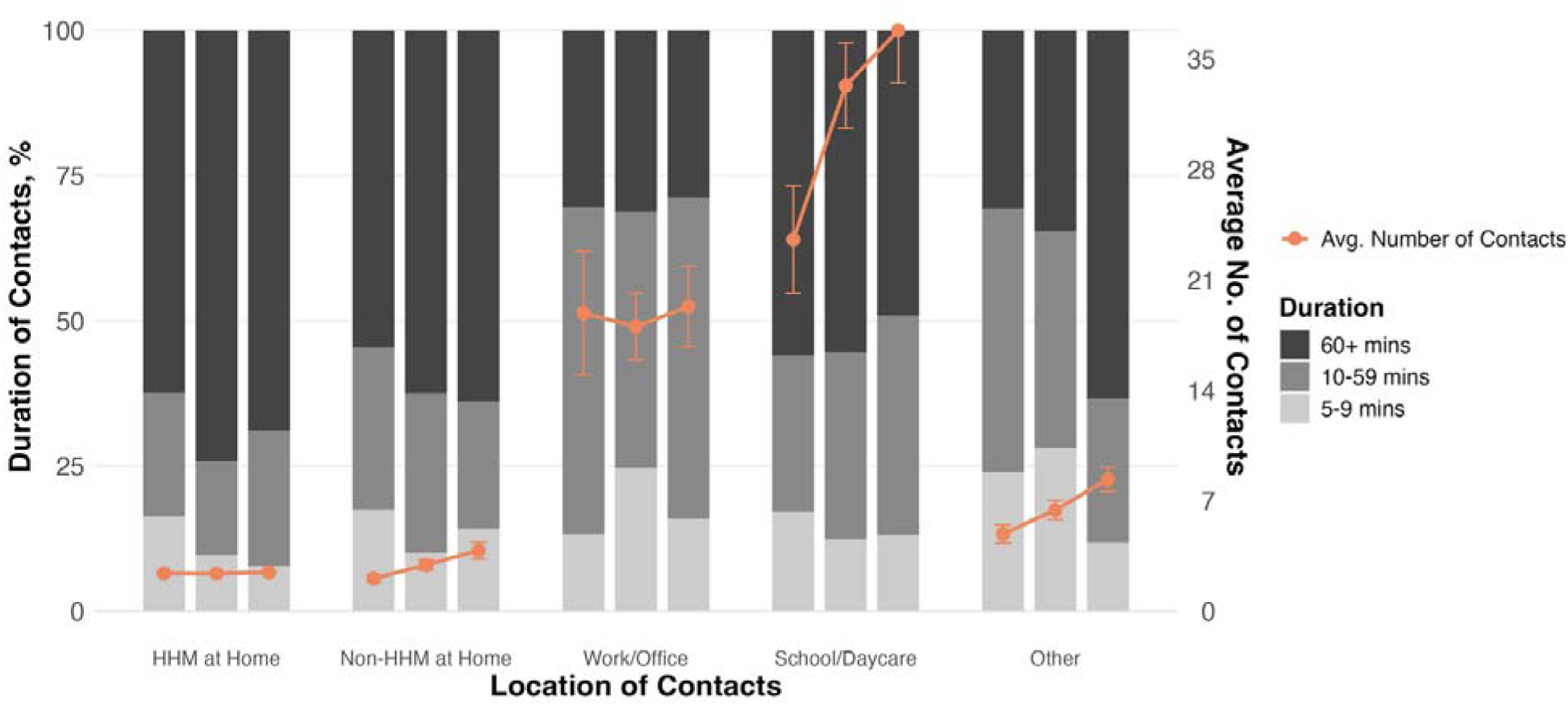
Durations and weighted average numbers of social contacts reported by acute gastroenteritis and acute respiratory infection cases over the course of illness by setting. Abbreviation: HHM: household members. The figure displays weighted mean contacts for cases from 500 bootstrap resampled iterations of N=1,000. Colored bars represent the proportion of social contact durations (5–9, 10–59, and 60+ minutes) (left axis). For each group of three bars, the left bar represents contacts made at index date, the middle bar represents contacts made at one-week follow-up and the right bar represents contacts made at two-week follow-up. The orange data points indicate the average number of social contacts in each setting over time, with vertical lines representing the standard deviation of the bootstrap means (right axis).

We assessed temporal changes in social contacts across various characteristics of AGE and ARI cases, including sex, racial group, ethnicity, household income, employment status, occupation, school attendance, residential setting, healthcare encounter type, and disease type (**Supplementary Figure 2**). Increases in social contacts over the course of illness were generally observed across many subgroups, but we also noted some heterogeneity. Contacts increased over time among White and African American cases but remained relatively stable among Asian cases. Contacts also increased among employed adult cases, while remaining relatively stable among those who were unemployed. Increases were observed among healthcare and education workers, with education workers reporting particularly high numbers of contacts even on the index date. Among those attending school, increases in contacts were observed among in-person attendees, whereas those attending remotely showed little change over time. When stratified by healthcare encounter type, social contacts increased over the course of illness among individuals who received virtual care or visited ambulatory clinics but remained low among those who visited the emergency department or were hospitalized. The number of days between the index date and the first survey completion did not meaningfully influence the overall temporal changes in social contacts (**Supplementary Figure 3**).

Social contacts with household members at home remained relatively stable over the course of illness across all age groups (**Supplementary Figure 4**). In contrast, social contacts with non-household members at work, school, or general community settings increased over time in most age groups. Social contacts with non-household members at home were infrequent, averaging fewer than one contact across all age groups, but also increased over the course of illness in most age groups.

### Household members of AGE and ARI cases

Of 923 household members that provided consent, 713 (77%) completed social contact surveys both on the index date and 1-week follow up. Of these, four cases were excluded from the analysis because their reported social contacts exceeded the threshold, resulting in a total of 709 household members included in the main analysis. Among 709 household members who completed all longitudinal social contact surveys, most were adults aged 30-44 years (31.5%) and children aged 5–17 years (27.8%) (**Supplementary Table 5**).

The mean number of social contacts among household members who completed all surveys remained relatively consistent over time (**Figure 1**), with 6.8 contacts (95% CI: 6.0–7.6) reported on the index date and 6.3 contacts (95% CI: 5.6–7.1) at the one-week follow-up. While individual data points showed substantial heterogeneity, distinct patterns emerged when household members were stratified by self-reported health status (**Supplementary Figure 5**).

The average number of contacts decreased among household members who were well on the index date but became ill during follow-up, whereas contacts increased among those who were also ill on the index date and had recovered by the one-week follow-up. The duration of social contacts reported by household members was relatively stable over time across settings (**Supplementary Figure 6**).

### Infection mitigation behaviors

The majority (80.2%) of 1,000 AGE and ARI cases that completed all surveys reported engaging in at least one mitigation behavior on the index date (**Table 1**). The most commonly reported mitigation behavior on the index date was more frequent handwashing or use of hand sanitizer (52.8%), followed by reducing the duration of contact (44.0%), increasing physical distance from others (43.9%), and missing work or school (43.8%). The proportion of cases reporting any mitigation behavior declined from 80.2% on the index date to 57.0% at the one-week follow-up and 36.0% at the two-week follow-up.

**Table 1.** Infection mitigation behaviors reported by acute gastroenteritis and acute respiratory infection cases who completed all longitudinal surveys over the course of acute infection (N=1,000).

| Mitigation behaviors | Index date | 1-week follow-up | 2-week follow-up |
|---|---|---|---|
|  | **N (95% CI), % (95% CI)** | **N (95% CI), % (95% CI)** | **N (95% CI), % (95% CI)** |
| Did not take any mitigation behaviors | 198 (173–221),<br>19.8% (17.3–22.1%) | 430 (397–456),<br>43.0% (39.7–45.6%) | 640 (609–670),<br>64.0% (60.9–67.0%) |
| Reduced contacts at home | 379 (348–410),<br>37.9% (34.8–41.0%) | 173 (150–200),<br>17.3% (15.0–20.0%) | 96 (77–114),<br>9.6% (7.7–11.4%) |
| Reduced contacts at work | 169 (146–191),<br>16.9% (14.6–19.1%) | 134 (112–153),<br>13.4% (11.2–15.3%) | 86 (69–105),<br>8.6% (6.9–10.5%) |
| Reduced contacts at school/daycare | 120 (100–140),<br>12.0% (10.0–14.0%) | 60 (45–76),<br>6.0% (4.5–7.6%) | 36 (24–46),<br>3.6% (2.4–4.6%) |
| Reduced contacts in the community | 407 (377–442),<br>40.7% (37.7–44.2%) | 224 (200–249),<br>22.4% (20.0–24.9%) | 94 (75–111),<br>9.4% (7.5–11.1%) |
| Reduced duration of interactions | 440 (409–472),<br>44.0% (40.9–47.2%) | 218 (192–244),<br>21.8% (19.2–24.4%) | 111 (93–129),<br>11.1% (9.3–12.9%) |
| Increased physical distance during interactions | 439 (408–471),<br>43.9% (40.8–47.1%) | 271 (244–298),<br>27.1% (24.4–29.8%) | 155 (132–178),<br>15.5% (13.2–17.8%) |
| Had contacts outside instead of inside | 40 (29–53),<br>4.0% (2.9–5.3%) | 65 (51–83),<br>6.5% (5.1–8.3%) | 46 (35–60),<br>4.6% (3.5–6.0%) |
| Wore a mask more frequently than usual | 372 (342–402),<br>37.2% (34.2–40.2%) | 203 (181–230),<br>20.3% (18.1–23.0%) | 111 (93–129),<br>11.1% (9.3–12.9%) |
| Cleaned hands more frequently | 528 (496–559),<br>52.8% (49.6–55.9%) | 414 (385–451),<br>41.4% (38.5–45.1%) | 277 (251–305),<br>27.7% (25.1–30.5%) |
| Missed work or school | 438 (409–471),<br>43.8% (40.9–47.1%) | 105 (88–126),<br>10.5% (8.8–12.6%) | 28 (19–38),<br>2.8% (1.9–3.8%) |

Abbreviation: CI: Confidence interval.
This table displays weighted estimates from 500 bootstrap resampled iterations of N=1,000.
Unweighted values can be found in Supplementary Table 7.

We assessed how self-reported mitigation behaviors corresponded to the reported number of social contacts. Cases who took mitigation behaviors that would reduce opportunities for social interaction (e.g., reduced home or community contacts, missed school/work) generally reported lower numbers of social contacts, compared to those who did not (**Supplementary Figure 7**). In contrast, behaviors that are intended to reduce transmission risks rather than limiting social interactions (e.g., more frequent hand washing or mask use) were not consistently associated with fewer contacts. Cases who did not take any mitigation behaviors reported a higher number of social contacts on the index date.

Among household members, 409 (57.7%) and 365 (51.5%) reported engaging in mitigation behaviors on the index date and 1-week follow up, respectively (**Supplementary Table 6**). Similar to cases, the most frequently reported mitigation behavior among household members was more frequent handwashing or use of hand sanitizer (39.6% on the index date and 33.7% at the one-week follow up).

### Sensitivity analysis

To evaluate the potential for attrition bias, we calculated summary statistics for social contacts among all cases who completed each corresponding survey, regardless of whether they completed all three surveys. Their reported social contacts were generally similar to those reported by cases who completed all three surveys. The unweighted mean number of contacts on the index date was 6.8 among all cases who completed the index-date survey, compared with 5.6 among those who completed all three surveys. At the one-week follow-up, the corresponding means were 9.7 and 9.5, respectively.

In a sensitivity analysis where we capped the total contacts per person at 50 among cases, temporal changes in social contact over the course of illness remained the same. The mean contacts at the 2-week follow-up was approximately double (1.92; 95% CI: 1.75–2.11) the mean contacts reported during the index survey.

Unweighted mean numbers of contacts for all characteristics examined in this study are presented in **Supplementary Figure 8**.

## Discussion

This study provides the U.S.-based longitudinal social contact data across all ages, capturing both symptomatic cases and their exposed close contacts, throughout the course of acute enteric and respiratory infections. These data were collected in the post-COVID-19 pandemic time, which may have altered public perceptions and behaviors surrounding infection mitigation. We found that symptomatic cases reduced their contacts at the peak of illness and gradually increased them over the following two weeks. Importantly, this increase was also observed among cases who reported that they remained sick at the one- and two-week follow-ups, potentially contributing to ongoing disease transmission. These findings underscore the importance of capturing realistic social contact behavior during acute infection in transmission models, especially when projecting outbreak trajectories and estimating intervention impacts, as is increasingly being done in the study of chronic infection and other sexually transmitted diseases (*24*, *25*).

Our participants generally reported a higher number of contacts than participants in previous social contact studies, even when they were sick. For example, a nationwide U.S. social contact study reported an average of 7.4 contacts per day (*32*). This difference was observed even though we asked respondents to report only contacts lasting more than five minutes, in an effort to reduce reporting burden and facilitate survey completion among participants who were actively ill. Differences in survey design may have contributed to the higher contact counts observed in our study. Traditional social contact diaries often ask participants to report information on each contact one by one, including characteristics such as age, sex, setting, and duration. In contrast, our survey only asked participants to report aggregate numbers of contacts by contact type, age group, and setting, which substantially reduced survey length and response time. This simplified approach may have enabled participants to report contacts that might otherwise have been omitted because of the burden associated with detailed contact diaries. In addition, we weighted our study population to represent medically attended AGE and ARI cases in the U.S., and the age distribution of our weighted study population may have also contributed to the higher contact counts. For example, cases aged 5–17 years (which generally have more social contacts) comprised 24% of our weighted sample, compared with 16% of the general U.S. population. These considerations suggest that absolute contact counts may be sensitive to survey design and the composition of the study population. Accordingly, the relative changes in social contacts over the course of illness may be more informative than the absolute contact counts themselves.

In examining temporal changes among symptomatic cases, our data showed a consistent pattern of reduced contacts on the index date followed by steady increases at one and two weeks later. This dynamic was particularly pronounced among school-aged children (5–17 years of age), who not only had the highest contact counts overall but also showed the largest increases over the course of infection. This pattern highlights the central role of school-aged children in social contact networks and their potential importance in driving disease transmission.

Our findings of reduced contacts during illness align more closely with the U.K. study of ARI cases, which also reported marked declines in contacts while sick (*26*). In contrast, the Malawi study found relatively stable total contact counts over the course of illness (*27*). This was explained by increased at-home contacts with non-household members during illness, which offset reductions in contacts outside the home. In Malawi, neighbors and extended family members commonly helped individuals during illness, a pattern that was not observed in our data or in the U.K. study. These contrasts highlight that illness-related changes in social behavior may differ substantially across cultural contexts, social structures, and baseline contact patterns. Although our sample size was insufficient to examine more detailed differences across racial or ethnic groups, future studies could explore whether illness-related contact changes also vary across subpopulations within the U.S. with different social and cultural practices.

Temporal changes also varied by setting in our data. Contact counts at home remained stable, while those outside the home decreased during illness, which was also observed in the U.K. (*26*). The number of social contacts among cases sharply increased at school and daycare settings over time. In contrast, work contacts showed no change at the aggregate level, although clear increases were observed among healthcare and education workers. This may reflect variation across occupational groups, as reported in studies from England (*33*) and the U.S. (*34*) during the COVID-19 pandemic. Education workers maintained higher contact levels even while ill, further highlighting the influence of occupation on contact patterns and disease transmission.

Our study provides unique data on social contact patterns among exposed close contacts of AGE and ARI cases. On average, their number of contacts remained stable over time, but patterns diverged when stratified by health status. Those who became ill tended to reduce their contacts, similar to what was observed among cases, whereas those who remained healthy showed substantial heterogeneity in their temporal changes. Social contact data from household members of acute infection cases are valuable, as these individuals are at elevated risk of infection. Even if they did not develop symptoms, they may have been asymptomatically infected and infectious without knowing. Household member data provide important insight into whether and how people adjust their social behaviors based on awareness of exposure, independent of their own symptomatic status.

Outcomes of mathematical models of infectious diseases are often sensitive to assumptions about human behavior, including who contacts whom and how contact patterns change over the course of infection. These assumptions may influence estimates of key model parameters as well as projected intervention effects.

Additionally, accounting for age-specific differences in behavioral changes may be important, particularly for groups with both high contact rates and substantial temporal changes in contact patterns, such as school-aged children. Models intended to project disease transmission or intervention impact should therefore consider incorporating illness-associated changes in social behavior when appropriate, and our publicly available contact data can support such efforts in future transmission modeling studies.

In addition to modeling applications, our findings also have practical implications for public health messaging and policy. The observed reduction in contacts while ill supports guidance that encourages people to stay home from work or school when experiencing acute symptoms. However, contacts were high even on the index date at school and work, and among cases that reported they were still ill at 1- and 2-week follow-ups. These insights underscore the importance of workplace leave policies, school absence policies, and clear communication about when it is safe to return to regular activities. Incorporating such behaviorally informed recommendations into public health strategies could help reduce transmission, particularly in high-contact settings such as schools.

Our study has several limitations. Our study population was restricted to members of KPNW, who may differ socioeconomically and demographically from the broader U.S. population, potentially limiting generalizability. We accounted for this through survey weighting based on age and sex using national data from MarketScan and Medicare. However, we were not able to adjust for other sociodemographic factors, such as income. We were also unable to weight the household member data because nationally representative data on the sociodemographic distribution of household members of AGE and ARI cases were not available. In addition, contact patterns were self-reported and therefore subject to recall bias and social desirability bias. We sought to minimize recall bias by excluding respondents whose index date was more than six days before survey completion, and in the final analytic sample, more than half of the participants (55.5%) reported an index date within three days of completing the first survey.

Importantly, when we compared temporal changes in social contacts between participants who completed the survey within three days of the index date versus those who did so within four to six days, we found no meaningful differences (**Supplementary Figure 3**). For the one- and two-week follow-up surveys, respondents were able to complete the surveys immediately after the corresponding follow-up dates, so there was less concern of recall bias. Of note, in excluding respondents whose index date was more than six days prior to survey completion, sicker patients may have been differentially excluded, either due to missing the survey completion deadline or due to deciding not to participate. To address social desirability bias, we noted in the survey that participants’ responses would not be shared with their healthcare providers or used in their medical care, and that their responses would not be judged. Also, because we recruited individuals who were actively ill, we minimized participant burden by simplifying and shortening the survey. As a result, we did not ask participants to distinguish between different types of social contacts (e.g., physical, verbal, or close-proximity) and asked for the total number of all social contacts combined. This approach was supported by findings from a previous study, which showed that transmission models for acute infections fit data better when all types of contacts were aggregated, compared to using only physical contacts (*26*). Nevertheless, asking participants to report aggregate numbers of contacts by time and setting may have introduced reporting error, leading some to over- or underestimate their contacts. In addition, some subgroups had sparse data, such as young children, which may limit the generalizability of their findings. Our study population was also predominantly White, highly educated, and from higher-income households, and these characteristics could not be fully accounted for through the available weighting data.

In conclusion, this study provides the first U.S.-based longitudinal data on social contact patterns during acute gastroenteritis and respiratory infections, capturing both symptomatic cases and their household members in the post-COVID-19 era. Our findings underscore the need to incorporate dynamic, illness-driven behavior changes into transmission models when projecting disease transmission and intervention impact. By making age-stratified contact matrices publicly available, this study provides critical empirical data for refining epidemic models and highlights the value of ongoing longitudinal surveillance to inform policies and interventions aimed at mitigating the spread of acute infectious diseases.

## Acknowledgments

This research was supported by the United States National Science Foundation (NSF) Incorporating Human Behavior in Epidemiological Models (IHBEM) grant (2327697). This study was also made possible by cooperative agreement CDC-RFA-FT-23-0069 from the CDC’s Center for Forecasting and Outbreak Analytics. The funders of the study had no role in study design, data collection, data analysis, data interpretation, or writing of the report.We thank study participants for sharing their data. We thank KPNW staff for data extraction and collection. We also thank Anushka Reddy Marri, Miwa Watanabe, and Richard Quach for their roles in survey testing and Machi Shiiba and Connor Van Meter for MarketScan data curation. The authors declare that they have no competing interests. Author contributions: Conceptualization: K.S., B.L., S.J. M.S. Methodology: K.S., B.L., S.J., M.S., M.L., D.C., A.S., J.I., J.D. Data Curation: J.D., M.S., A.T., G.L., G.L., J.I., K.S. Formal Analysis: J.I., G.L. G.L., A.T., K.S. Validation: J.I., K.S., A.S., B.L., M.S., S.J., M.L., J.D. Writing: K.S., J.I., G.L., G.L., A.T. Reviewing & Editing: All authors. Supervision: K.S., B.L., M.S. Funding Acquisition: K.S., B.L., S.J., M.S.

## Data Availability

Individual-level participant data cannot be shared publicly because public release is not permitted under the study’s approved consent and data-sharing framework. To maximize transparency and reproducibility within these restrictions, we have made the most granular data that can be publicly shared available at https://github.com/KayokoShioda/SickMix. These materials include aggregated age-stratified social contact matrices for the index date and the one-week and two-week follow-ups, as well as the relevant analysis code, documentation, variable definitions, and aggregate inputs used for the manuscript analyses.

## References

1. N. Bharti, Linking human behaviors and infectious diseases. Proc. Natl. Acad. Sci. U. S. A. 118, e2101345118 (2021).

2. C. Buckee, A. Noor, L. Sattenspiel, Thinking clearly about social aspects of infectious disease transmission. Nature 595, 205–213 (2021).

3. S. Bansal, B. T. Grenfell, L. A. Meyers, When individual behaviour matters: homogeneous and network models in epidemiology. J. R. Soc. Interface 4, 879–891 (2007).

4. S. Funk, M. Salathé, V. A. A. Jansen, Modelling the influence of human behaviour on the spread of infectious diseases: a review. J. R. Soc. Interface 7, 1247–1256 (2010).

5. F. Verelst, L. Willem, P. Beutels, Behavioural change models for infectious disease transmission: a systematic review (2010–2015). J. R. Soc. Interface 13, 20160820 (2016).

6. N. Perra, D. Balcan, B. Gonçalves, A. Vespignani, Towards a characterization of behavior-disease models. PloS One 6, e23084 (2011).

7. J. Mossong, N. Hens, M. Jit, P. Beutels, K. Auranen, R. Mikolajczyk, M. Massari, S. Salmaso, G. S. Tomba, J. Wallinga, J. Heijne, M. Sadkowska-Todys, M. Rosinska, W. J. Edmunds, Social Contacts and Mixing Patterns Relevant to the Spread of Infectious Diseases. PLOS Med. 5, e74 (2008).

8. M. Ajelli, M. Litvinova, Estimating contact patterns relevant to the spread of infectious diseases in Russia. J. Theor. Biol. 419, 1–7 (2017).

9. J. C. Blackwood, D. A. T. Cummings, S. Iamsirithaworn, P. Rohani, Using age-stratified incidence data to examine the transmission consequences of pertussis vaccination. Epidemics 16, 1–7 (2016).

10. P. T. Campbell, J. McVernon, N. Shrestha, P. M. Nathan, N. Geard, Who’s holding the baby? A prospective diary study of the contact patterns of mothers with an infant. BMC Infect. Dis. 17, 634 (2017).

11. Y. Fu, D.-W. Wang, J.-H. Chuang, Representative Contact Diaries for Modeling the Spread of Infectious Diseases in Taiwan. PLoS ONE 7, e45113 (2012).

12. C. G. Grijalva, N. Goeyvaerts, H. Verastegui, K. M. Edwards, A. I. Gil, C. F. Lanata, N. Hens, for the RESPIRA PERU project, A Household-Based Study of Contact Networks Relevant for the Spread of Infectious Diseases in the Highlands of Peru. PLOS ONE 10, e0118457 (2015).

13. Y. Ibuka, Y. Ohkusa, T. Sugawara, G. B. Chapman, D. Yamin, K. E. Atkins, K. Taniguchi, N. Okabe, A. P. Galvani, Social contacts, vaccination decisions and influenza in Japan. J. Epidemiol. Community Health 70, 162–167 (2016).

14. S. P. Johnstone-Robertson, D. Mark, C. Morrow, K. Middelkoop, M. Chiswell, L. D. H. Aquino, L.-G. Bekker, R. Wood, Social Mixing Patterns Within a South African Township Community: Implications for Respiratory Disease Transmission and Control. Am. J. Epidemiol. 174, 1246–1255 (2011).

15. M. C. Kiti, T. M. Kinyanjui, D. C. Koech, P. K. Munywoki, G. F. Medley, D. J. Nokes, Quantifying Age-Related Rates of Social Contact Using Diaries in a Rural Coastal Population of Kenya. PLoS ONE 9, e104786 (2014).

16. G. M. Knight, N. J. Dharan, G. J. Fox, N. Stennis, A. Zwerling, R. Khurana, D. W. Dowdy, Bridging the gap between evidence and policy for infectious diseases: How models can aid public health decision-making. Int. J. Infect. Dis. 42, 17–23 (2016).

17. C. Doran, M. Sheku, Y. Nakada, B. Lopman, K. Nelson, Relevance of social contact definitions for use in infectious disease transmission modeling: a systematic review and recommendations. BMC Infect. Dis. 26, 836 (2026).

18. S. Zissette, M. C. Kiti, B. W. Bennett, C. Y. Liu, K. N. Nelson, A. Zelaya, J. T. Kellogg, T. M. Johnson Ii, P. Clayton, S. K. Fridkin, S. B. Omer, B. A. Lopman, C. Adams, Social contact patterns among employees in U.S. long-term care facilities during the COVID-19 pandemic, December 2020 to June 2021. BMC Res. Notes 16, 294 (2023).

19. D. M. Feehan, A. S. Mahmud, Quantifying population contact patterns in the United States during the COVID-19 pandemic. Nat. Commun. 12, 893 (2021).

20. M. C. Kiti, O. G. Aguolu, A. Zelaya, H. Y. Chen, N. Ahmed, J. Batross, C. Y. Liu, K. N. Nelson, S. M. Jenness, A. Melegaro, F. Ahmed, F. Malik, S. B. Omer, B. A. Lopman, Changing social contact patterns among US workers during the COVID-19 pandemic: April 2020 to December 2021. Epidemics 45, 100727 (2023).

21. C. Y. Liu, J. Berlin, M. C. Kiti, E. Del Fava, A. Grow, E. Zagheni, A. Melegaro, S. M. Jenness, S. B. Omer, B. Lopman, K. Nelson, Rapid Review of Social Contact Patterns During the COVID-19 Pandemic. Epidemiology 32, 781–791 (2021).

22. M. J. Kozal, K. R. Amico, J. Chiarella, D. Cornman, W. Fisher, J. Fisher, G. Friedland, A PopulationDBased and Longitudinal Study of Sexual Behavior and MultidrugDResistant HIV Among Patients in Clinical Care. J. Int. AIDS Soc. 8, 72–72 (2006).

23. I. W. G. A. E. Putra, N. M. D. Kurniasari, N. P. E. P. Dewi, I. K. Suarjana, I. M. K. Duana, I. K. H. Mulyawan, P. Riono, B. Alisjahbana, A. Probandari, H. B. Notobroto, C. U. Wahyuni, The Implementation of Early Detection in Tuberculosis Contact Investigation to Improve Case Finding. J. Epidemiol. Glob. Health, doi: 10.2991/jegh.k.190808.001 (2019).

24. X. Sun, W. Zhou, Y. Ruan, G. Lan, Q. Zhu, Y. Xiao, Perceived risk induced multiscale model: Coupled within-host and between-host dynamics and behavioral dynamics. J. Theor. Biol. 599, 111998 (2025).

25. M. A. L. Hayashi, M. C. Eisenberg, Effects of adaptive protective behavior on the dynamics of sexually transmitted infections. J. Theor. Biol. 388, 119–130 (2016).

26. K. Van Kerckhove, N. Hens, W. J. Edmunds, K. T. D. Eames, The impact of illness on social networks: implications for transmission and control of influenza. Am. J. Epidemiol. 178, 1655–1662 (2013).

27. J. R. Glynn, E. McLean, J. Malava, A. Dube, C. Katundu, A. C. Crampin, S. Geis, Effect of Acute Illness on Contact Patterns, Malawi, 2017. Emerg. Infect. Dis. 26, 44–50 (2020).

28. M. A. Schmidt, H. C. Groom, A. L. Naleway, C. Biggs, S. B. Salas, K. Shioda, Z. Marsh, J. L. Donald, A. J. Hall, A model for rapid, active surveillance for medically-attended acute gastroenteritis within an integrated health care delivery system. PLOS ONE 13, e0201805 (2018).

29. D. J. A. Toth, M. Leecaster, W. B. P. Pettey, A. V. Gundlapalli, H. Gao, J. J. Rainey, A. Uzicanin, M. H. Samore, The role of heterogeneity in contact timing and duration in network models of influenza spread in schools. J. R. Soc. Interface 12, 20150279 (2015).

30. G. E. Potter, M. S. Handcock, I. M. Longini, M. E. Halloran, ESTIMATING WITHIN-SCHOOL CONTACT NETWORKS TO UNDERSTAND INFLUENZA TRANSMISSION. Ann. Appl. Stat. 6, 1–26 (2012).

31. T. Smieszek, S. Castell, A. Barrat, C. Cattuto, P. J. White, G. Krause, Contact diaries versus wearable proximity sensors in measuring contact patterns at a conference: method comparison and participants’ attitudes. BMC Infect. Dis. 16, 341 (2016).

32. M. Litvinova, S. Sinclair, A. G. Kummer, P. C. Ventura, T. Foster, K. Shioda, M. E. Halloran, A. Vespignani, M. Ajelli, Epistorm-Mix: Mapping Social Contact Patterns for Respiratory Pathogen Spread in the Post-Pandemic United States. medRxiv [Preprint] (2025). 10.1101/2025.11.20.25340662.

33. S. Beale, S. Hoskins, T. Byrne, W. L. E. Fong, E. Fragaszy, C. Geismar, J. Kovar, A. M. D. Navaratnam, V. Nguyen, P. Patel, A. Yavlinsky, A. M. Johnson, M. V. Tongeren, R. W. Aldridge, A. Hayward, Workplace contact patterns in England during the COVID-19 pandemic: Analysis of the Virus Watch prospective cohort study. Lancet Reg. Health – Eur. 16 (2022).

34. K. N. Nelson, A. J. Siegler, P. S. Sullivan, H. Bradley, E. Hall, N. Luisi, P. Hipp-Ramsey, T. Sanchez, K. Shioda, B. A. Lopman, Nationally representative social contact patterns among U.S. adults, August 2020-April 2021. Epidemics 40, 100605 (2022).

