## Supplementary Material for "SickMix: Temporal Changes in Social Contact Patterns among People with Acute Infection and Their Close Contacts"

Jessica C. Ibiebele^1^, Aarushi Tuli^2^*, Grissel Lopes^2^*, Gina Lombard^2^*, Anne Shapiro^3^, Judy Donald^4^, Mark A. Schmidt ^4^, Maria Litvinova^5^, Dehao Chen^6^, Samuel M. Jenness^6^, Benjamin A. Lopman^6^, Kayoko Shioda^1,7^

^1^Center on Emerging Infectious Diseases, Boston University, Boston, MA, USA

^2^School of Public Health, Boston University, Boston, MA, USA

^3^Department of Biostatistics, School of Public Health, Boston University, Boston, MA, USA

^4^Center for Health Research, Kaiser Permanente Northwest, Portland, OR, USA

^5^Department of Epidemiology and Biostatistics, School of Public Health, Indiana University, Bloomington, IN, USA

^6^Department of Epidemiology, Rollins School of Public Health, Emory University, Atlanta, GA, USA

^7^Department of Global Health, School of Public Health, Boston University, Boston, MA, USA

*Co-second authors

Corresponding Author:

Kayoko Shioda, PhD, DVM, MPH

Department of Global Health, School of Public Health, Boston University, Boston, MA;

Center on Emerging Infectious Diseases, Boston University, Boston, MA

(617) 358-2422

### Supplementary Tables

#### Supplementary Table 1. ICD-10 codes for acute respiratory infections.

| **Diagnosis** | **ICD-10** |
| --- | --- |
| **Acute Upper Respiratory Infections** | J00-J06 |
| Acute nasopharyngitis [common cold] | J00 |
| Acute sinusitis | J01.0- J01.9 |
| Acute pharyngitis | J02.0-J02.9 |
| Acute tonsillitis | J03.0-J03.9 |
| Acute laryngitis and tracheitis | J04.0-J04.3 |
| Acute obstructive laryngitis [croup] and epiglottitis | J05.0-J05.1 |
| Acute upper respiratory infections of multiple and unspecified sites | J06.0-J06.9 |
| **Influenza and Pneumonia** | J09-J18 |
| Influenza due to certain identified influenza viruses | J09.X-J09.X9 |
| Influenza due to other identified influenza virus | J10.0-J10.8 |
| Influenza due to unidentified influenza virus | J11.0-J11.8 |
| Viral pneumonia, not elsewhere classified | J12.0-J12.9 |
| Pneumonia due to Streptococcus pneumoniae | J13 |
| Pneumonia due to Hemophilus influenzae | J14 |
| Bacterial pneumonia, not elsewhere classified | J15.0-J15.9 |
| Pneumonia due to other infectious organisms, not elsewhere classified | J16.0-J16.8 |
| Pneumonia in diseases classified elsewhere | J17 |
| Pneumonia, unspecified organism | J18.0-J18.9 |

Abbreviation: ICD, International Classification of Diseases.

#### Supplementary Table 2. ICD-10 codes for acute gastroenteritis.

| **Diagnosis** | **ICD-10** |
| --- | --- |
| Infectious gastroenteritis and colitis, unspecified | A09 |
| Intestinal infectious diseases | A00-A09 |
| Noninfective gastroenteritis and colitis, unspecified | K52.9 |
| Rotaviral enteritis | A08.0-A08.5 |
| Cholera due to Vibrio cholerae 01, biovar cholerae | A00.0 |
| Bacterial foodborne intoxication, unspecified | A05.9 |
| Acute amebic dysentery | A06.0 |
| Protozoal intestinal disease, unspecified | A07.9 |

Abbreviation: ICD, International Classification of Diseases.

#### Supplementary Table 3. Characteristics of consented acute gastroenteritis and acute respiratory infection cases.

| **Characteristics** | **Analytic sample* (N=1,000)^1^, n, %** | **All enrolled cases (N=2,570)^1^, n, %** | **p-value^2^** |
| --- | --- | --- | --- |
| **Sex assigned at birth** |  |  |  |
| Female | 658, 65.8% | 1643, 63.9% | 0.2 |
| Male | 342, 34.2% | 927, 36.1% |  |
| **Age group (in years)** |  |  |  |
| <1 | 14, 1.4% | 39, 1.5% | 0.15 |
| 1-4 | 65, 6.5% | 140, 5.4% |  |
| 5-17 | 125, 12.5% | 291, 11.3% |  |
| 18-29 | 100, 10.0% | 302, 11.8% |  |
| 30-44 | 247, 24.7% | 573, 22.3% |  |
| 45-59 | 176 17.6% | 464, 18.1% |  |
| 60-74 | 182, 18.2% | 495, 19.3% |  |
| 75+ | 91, 9.1% | 266, 10.4% |  |

Abbreviations: AGE: Acute gastroenteritis, ARI: Acute respiratory infection.

*Includes everyone who completed 3 surveys and did not report excessive contacts

^1^n(%)

^2^Chi-squared goodness-of-fit test

#### Supplementary Table 4. Characteristics of acute gastroenteritis and acute respiratory infection cases who completed all longitudinal online social contact surveys.

| **Characteristics** | **Unweighted (N=1,000),**  **n, %** | **Weighted (N=1,000),**  **n (95% CI), % (95% CI)** |
| --- | --- | --- |
| **Diagnosis type** |  |  |
| Acute respiratory infection | 723, 72.3% | 991 (984–996), 99.1% (98.4–99.6%) |
| Acute gastroenteritis | 277, 27.7% | 9 (4–16), 0.9% (0.4–1.6%) |
| **Age group (in years)** |  |  |
| <1 | 14, 1.4% | 19 (11–28), 1.9% (1.1--2.8%) |
| 1–4 | 65, 6.5% | 89 (71–106), 8.9% (7.1–10.6%) |
| 5–17 | 125, 12.5% | 241 (217–271), 24.1% (21.7–27.1%) |
| 18–29 | 100,10.0% | 156 (136–180), 15.6% (13.6–18.0%) |
| 30–44 | 247, 24.7% | 214 (190–240), 21.4% (19.0–24.0%) |
| 45–59 | 176, 17.6% | 186 (163–211), 18.6% (16.3–21.1%) |
| 60–74 | 182, 18.2% | 76 (61–92), 7.6% (6.1–9.2%) |
| 75+ | 91, 9.1% | 17 (10–26), 1.7% (1.0–2.6%) |
| **Sex assigned at birth** |  |  |
| Female | 658, 65.8% | 586 (556–615), 58.6% (55.6–61.5%) |
| Male | 342, 34.2% | 414 (385–444), 41.4% (38.5–44.4%) |
| **Sexual orientation** |  |  |
| Heterosexual (straight) | 643, 64.3% | 497 (465–527), 49.7% (46.5–52.7%) |
| Lesbian | 20, 2.0% | 13 (6–20), 1.3% (0.6–2.0%) |
| Gay | 18, 1.8% | 18 (10–26), 1.8% (1.0–2.6%) |
| Bisexual | 68, 6.8% | 71 (55–87), 7.1% (5.5–8.7%) |
| Not sure/Questioning | 2, 0.2% | 1 (0–3), 0.1% (0.0–0.3%) |
| Something else | 12, 1.2% | 13 (7–21), 1.3% (0.7–2.1%) |
| Unknown/Declined to answer | 237, 23.7% | 387 (357–418), 38.7% (35.7–41.8%) |
| **Race** |  |  |
| American Indian or Alaska Native | 5, 0.5% | 7 (3–13), 0.7% (0.3–1.3%) |
| Asian | 50, 5.0% | 54 (41–68), 5.4% (4.1–6.8%) |
| Black or African American | 19, 1.9% | 22 (13–32), 2.2% (1.3–3.2%) |
| Native Hawaiian or other Pacific Islander | 3, 0.3% | 6 (2–11), 0.6% (0.2–1.1%) |
| White | 819, 81.9% | 762 (735–790), 76.2% (73.5–79.0%) |
| Other | 23, 2.3% | 32 (22–43), 3.2% (2.2–4.3%) |
| Multirace | 48, 4.8% | 77 (62–94), 7.7% (6.2–9.4%) |
| Unknown/Declined to Answer | 33, 3.3% | 38 (27–51), 3.8% (2.7–5.1%) |
| **Ethnicity** |  |  |
| Hispanic | 108, 10.8% | 138 (118–161), 13.8% (11.8–16.1%) |
| Non-Hispanic | 861, 86.1% | 821 (795–843), 82.1% (79.5–84.3%) |
| Unknown/Declined to answer | 31, 3.1% | 40 (29–53), 4.0% (2.9–5.3%) |
| **Residential setting** |  |  |
| House | 801, 80.1% | 808 (782–832), 80.8% (78.2–83.2%) |
| Condo or apartment | 191, 19.1% | 186 (162–214), 18.6% (16.2–21.4%) |
| Dormitory or other group setting such as military barracks | 3, 0.3% | 3 (0–7), 0.3% (0.0–0.7%) |
| Other | 4, 0.4% | 2 (0–5), 0.2% (0.0–0.5%) |
| Unknown/Declined to answer | 1, 0.1% | 1 (0–3), 0.1% (0.0–0.3%) |
| **Employment status** |  |  |
| Employed | 519, 51.9% | 508 (479–537), 50.8% (47.9–53.7%) |
| Not employed | 272, 27.2% | 135 (114–156), 13.5% (11.4–15.6%) |
| Unknown/ Declined to answer/Not applicable | 209, 20.9% | 356 (329–387), 35.6% (32.9–38.7%) |
| **Work setting** |  |  |
| Mostly outside the home | 324, 32.4% | 330 (302–360), 33.0% (30.2–36.0%) |
| Mostly inside the home | 104, 10.4% | 84 (68–101), 8.4% (6.8–10.1%) |
| Both outside of and inside the home (hybrid) | 91, 9.1% | 93 (74–112), 9.3% (7.4–11.2%) |
| Not applicable | 481, 48.1% | 492 (463–521), 49.2% (46.3–52.1%) |
| **Occupation** |  |  |
| Healthcare | 104, 10.4% | 88 (72–105), 8.8% (7.2–10.5%) |
| Food service | 14, 1.4% | 16 (10–24), 1.6% (1.0–2.4%) |
| Education | 137, 13.7% | 140 (119–159), 14.0% (11.9–15.9%) |
| Other | 264, 26.4% | 265 (236–292), 26.5% (23.6–29.2%) |
| Not applicable/Not reported | 481, 48.1% | 492 (463–521), 49.2% (46.3–52.1%) |
| **School attendance** |  |  |
| Yes | 215, 21.5% | 358 (329–389), 35.8% (32.9–38.9%) |
| Attend classes in person | 185, 18.5% | 320 (293–349), 32.0% (29.3–34.9%) |
| Do not attend classes in person | 30, 3.0% | 39 (28–52), 3.9% (2.8–5.2%) |
| No | 784, 78.4% | 642 (609–669), 64.2% (60.9–66.9%) |
| Unknown/Not sure | 1, 0.1% | 1 (0–4), 0.1% (0.0–0.4%) |
| **Highest level of education completed** |  |  |
| Less than high school | 7, 0.7% | 9 (4–17), 0.9% (0.4–1.7%) |
| High school or equivalent | 49, 4.9% | 51 (39–66), 5.1% (3.9–6.6%) |
| Vocational or trade school | 32, 3.2% | 23 (14–33), 2.3% (1.4–3.3%) |
| Some college | 140, 14.0% | 102 (84–121), 10.2% (8.4–12.1%) |
| College graduate | 265, 26.5% | 239 (209–263), 23.9% (20.9–26.3%) |
| Post-graduate | 299, 29.9% | 225 (199–252), 22.5% (19.9–25.2%) |
| Declined to answer/Not applicable | 208, 20.8% | 351 (325–382), 35.1% (32.5–38.2%) |
| **Total household income the past year (before taxes)** |  |  |
| Less than $15,000 | 24, 2.4% | 27 (18–38), 2.7% (1.8–3.8%) |
| Between $15,000 and $24,999 | 37, 3.7% | 29 (20–42), 2.9% (2.0–4.2%) |
| Between $25,000 and $49,999 | 95, 9.5% | 79 (63–95), 7.9% (6.3–9.5%) |
| Between $50,000 and $74,999 | 153, 15.3% | 144 (121–165), 14.4% (12.1–16.5%) |
| Between $75,000 and $99,999 | 150, 15.0% | 140 (120–161), 14.1% (12.0–16.1%) |
| Between $100,000 and $124,999 | 135, 13.5% | 135 (115–158), 13.5% (11.5–15.8%) |
| Between $125,000 and $149,999 | 105, 10.5% | 102 (82–121), 10.2% (8.2–12.1%) |
| Over $150,000 | 229, 22.9% | 279 (248–307), 27.9% (24.8–30.7%) |
| Missing | 72, 7.2% | 64 (49–83), 6.4% (4.9–8.3%) |
| **Healthcare encounter type** |  |  |
| Virtual care | 398, 39.8% | 320 (290–347), 32.0% (29.0–34.7%) |
| Ambulatory visit | 554, 55.4% | 660 (632–689), 66.0% (63.2–68.9%) |
| Emergency department | 46, 4.6% | 19 (12–28), 1.9% (1.2–2.8%) |
| Acute inpatient hospital stay | 2, 0.2% | 0 (0–2), 0.0% (0.0–0.2%) |
| **Index date*** |  |  |
| ≤3 days from the date of the enrollment survey | 566, 56.6% | 555 (521–584), 55.5% (52.1–58.4%) |
| 4-6 days from the date of the enrollment survey | 434, 43.4% | 445 (416–479), 44.5% (41.6–47.9%) |

Abbreviation: CI, confidence interval.

*Index date is the date cases felt most sick. The unweighted column contains observed data (N=1,000). The weighted column contains weighted estimates from 500 bootstrap resampled iterations of N=1,000.

#### Supplementary Table 5. Characteristics of household members that completed all longitudinal online social contact surveys (N=709).

| **Characteristics** | **Frequency (%)** |
| --- | --- |
| **Age group (in years)** |  |
| <5 | 83, 11.7% |
| 5-17 | 197, 27.8% |
| 18-29 | 27, 3.8% |
| 30-44 | 223, 31.5% |
| 45-59 | 87, 12.3% |
| 60-74 | 59, 8.3% |
| 75+ | 33, 4.7% |
| **Sex** |  |
| Female | 385, 54.3% |
| Male | 316, 44.6% |
| Non-binary | 8, 1.1% |
| **Disease type of the case in the household** |  |
| AGE | 175, 24.7% |
| ARI | 534, 75.3% |

Abbreviations: AGE: Acute gastroenteritis, ARI: Acute respiratory infection.

#### Supplementary Table 6. Infection mitigation behaviors reported by household members who completed all longitudinal surveys at the index date and 1-week follow-up (N=709).

| **Mitigation behaviors** | **Index date** | **1-week follow-up** |
| --- | --- | --- |
| Did not take any mitigation behaviors | 300, 42.3% | 344, 48.5% |
| Reduced contacts at home | 112, 15.8% | 75, 10.6% |
| Reduced contacts at work | 62, 8.7% | 47, 6.6%% |
| Reduced contacts at school/daycare | 40, 5.6% | 47, 6.6% |
| Reduced contacts in the community | 143, 20.2% | 102, 14.4% |
| Reduced duration of interactions | 147, 20.7% | 107, 15.1% |
| Increased physical distance during interactions | 154, 21.7% | 108, 15.2% |
| Had contacts outside instead of inside | 29, 4.1% | 26, 3.7% |
| Wore a mask more frequently than usual | 100, 14.1% | 75, 10.6% |
| Cleaned hands more frequently | 281, 39.6% | 239, 33.7% |
| Missed work or school | 80, 11.3% | 50, 7.1% |

#### Supplementary Table 7. Infection mitigation behaviors reported by acute gastroenteritis and acute respiratory infection cases who completed all longitudinal surveys over the course of acute infection, unweighted (N=1,000).

| **Mitigation behaviors** | **Index date** | **1-week follow-up** | **2-week follow-up** |
| --- | --- | --- | --- |
| Did not take any mitigation behaviors | 167, 16.7% | 395, 39.5% | 579, 57.9% |
| Reduced contacts at home | 382, 38.2% | 196, 19.6% | 111, 11.1% |
| Reduced contacts at work | 164, 16.4% | 132, 13.2% | 78, 7.8% |
| Reduced contacts at school/daycare | 93, 9.3% | 57, 5.7% | 33, 3.3% |
| Reduced contacts in the community | 395, 39.5% | 250, 25.0% | 126, 12.6% |
| Reduced duration of interactions | 432, 43.2% | 235, 23.5% | 137, 13.7% |
| Increased physical distance during interactions | 430, 43.0% | 277, 27.7% | 164, 16.4% |
| Had contacts outside instead of inside | 46, 4.6% | 73, 7.3% | 49, 4.9% |
| Wore a mask more frequently than usual | 351, 35.1% | 209, 20.9% | 120, 12.0% |
| Cleaned hands more frequently | 550, 55.0% | 433, 43.3% | 298, 29.8% |
| Missed work or school | 364, 36.4% | 103, 10.3% | 34, 3.4% |

### Supplementary Figures

#### Supplementary Figure 1. Weighted number of social contacts reported by acute gastroenteritis and acute respiratory infection cases over time by self-reported health status.

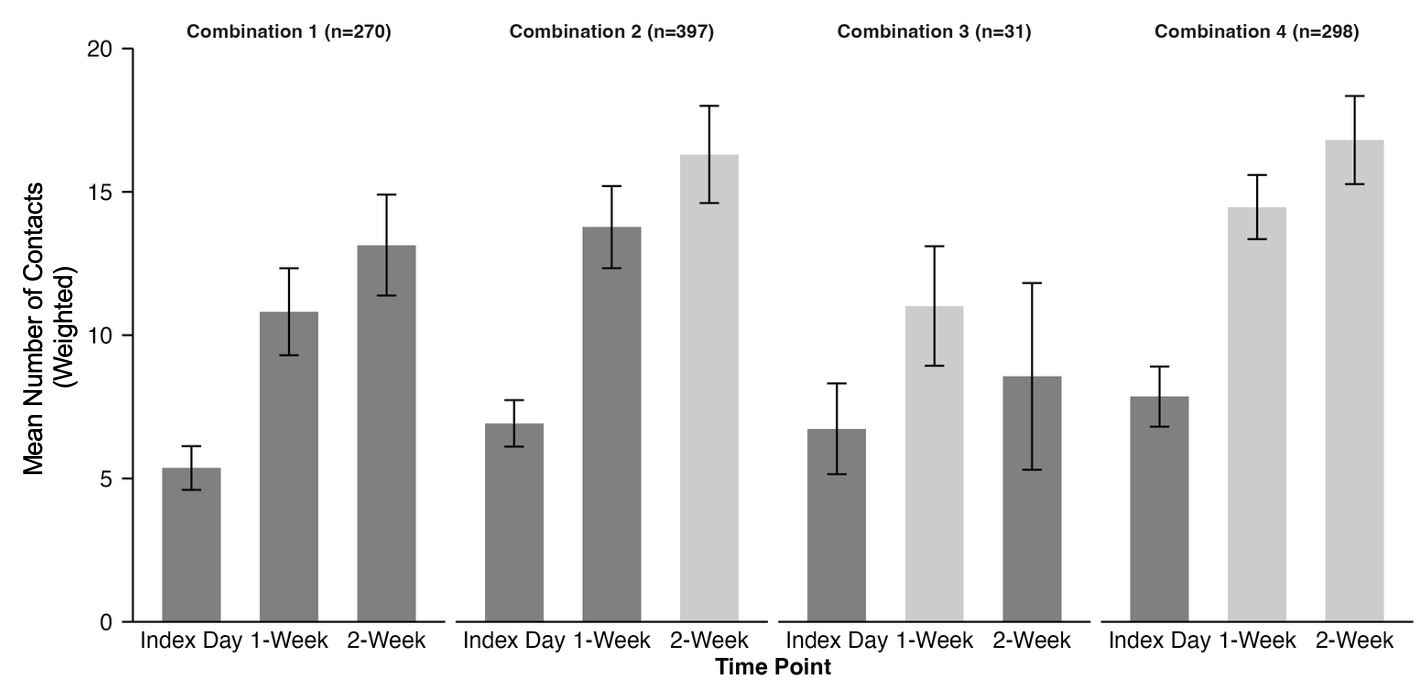

This figure presents the distribution of the number of contacts reported by acute gastroenteritis and acute respiratory infection cases on the index date, 1-Week-, and 2-Week Follow-Up, from 500 bootstrap resampled iterations of N=1,000 stratified into the following four mutually exclusive health status combinations:

- Combination 1 includes cases who reported feeling sick on all 3 days.
- Combination 2 includes those who felt sick on the index date and 1-Week Follow-Up but well at the 2-Week Follow-Up.
- Combination 3 includes those who felt sick on the index date, well at the 1-Week Follow-Up, and sick again at the 2-Week Follow-Up.
- Combination 4 includes those who were sick on the index date, but reported feeling well at the 1- and 2-Week Follow-Up.

Each combination displays three bars–one for each time point–showing the mean contacts and vertical lines representing the standard deviation of the bootstrap means. Bars in darker grey represent data when cases felt sick, and those in lighter grey represent data when cases felt well.

#### Supplementary Figure 2. Weighted mean number of social contacts reported by acute gastroenteritis and acute respiratory infection cases over the course of acute infection by characteristics and settings.

A. Sex	B. Racial group

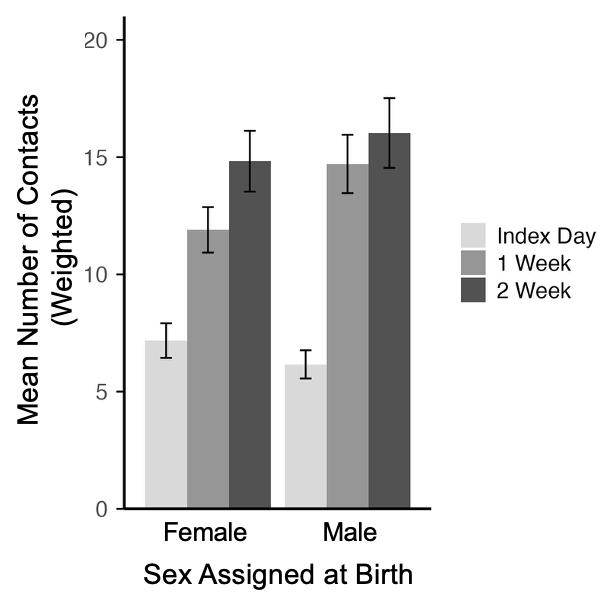

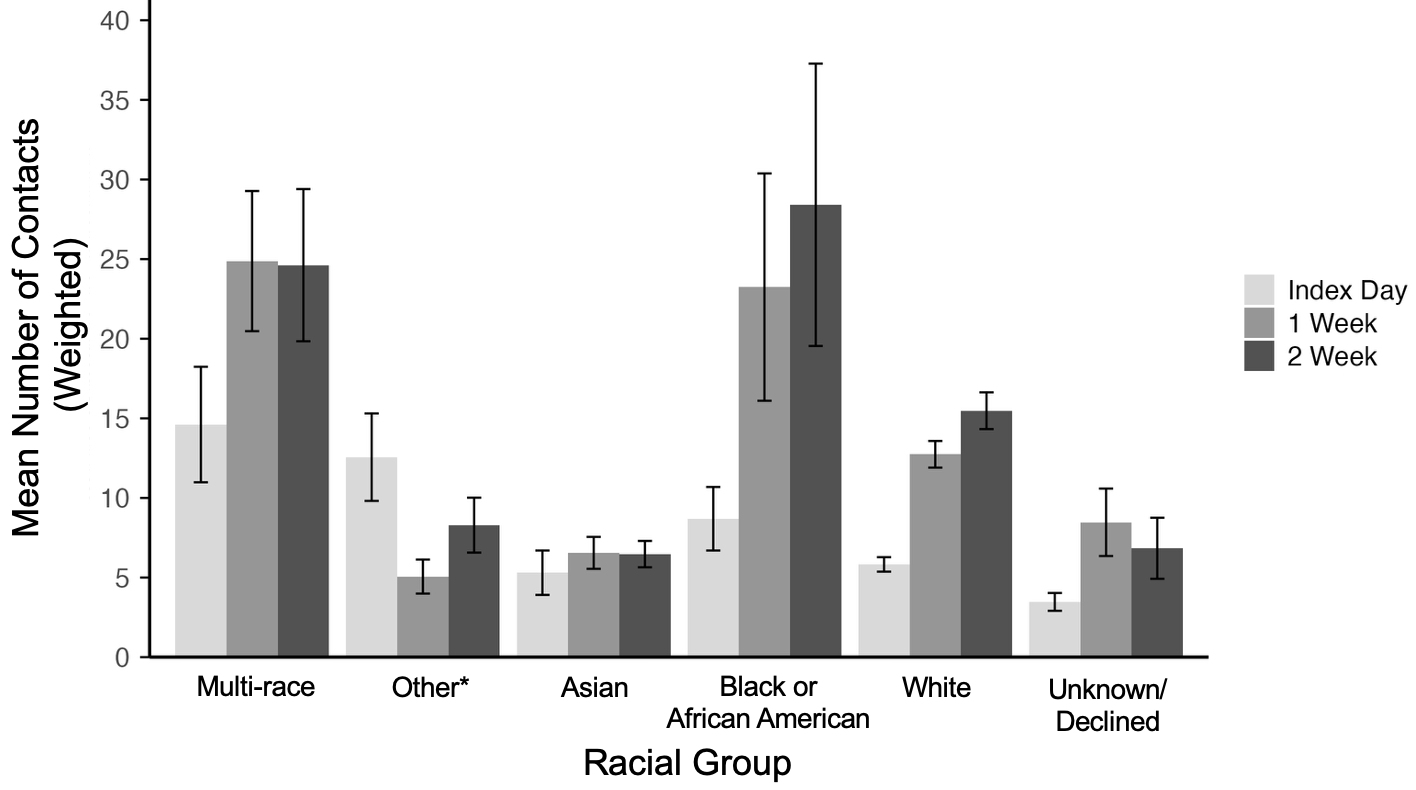

C. Ethnicity	D. Employment status

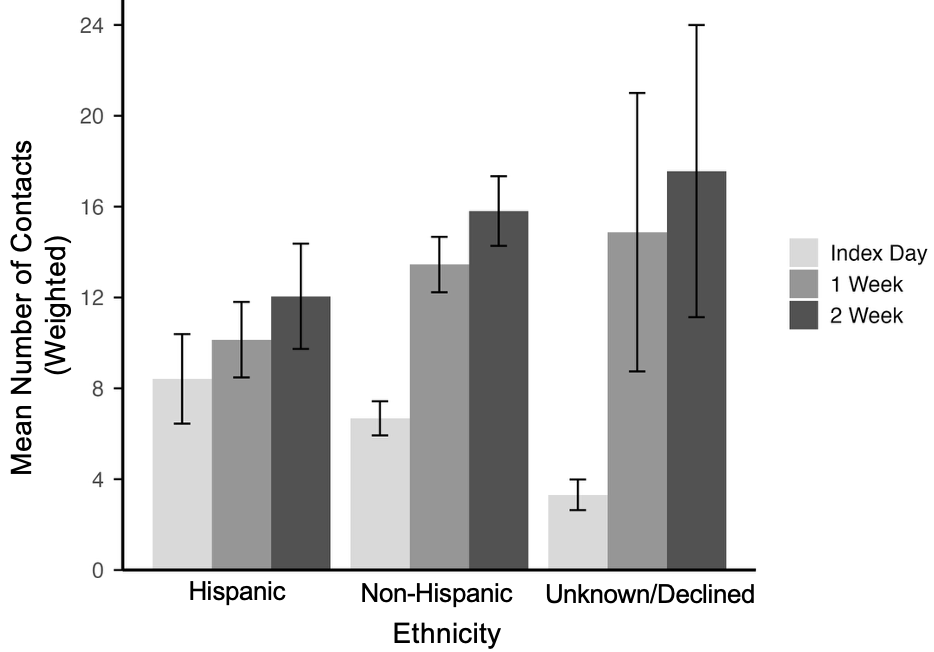

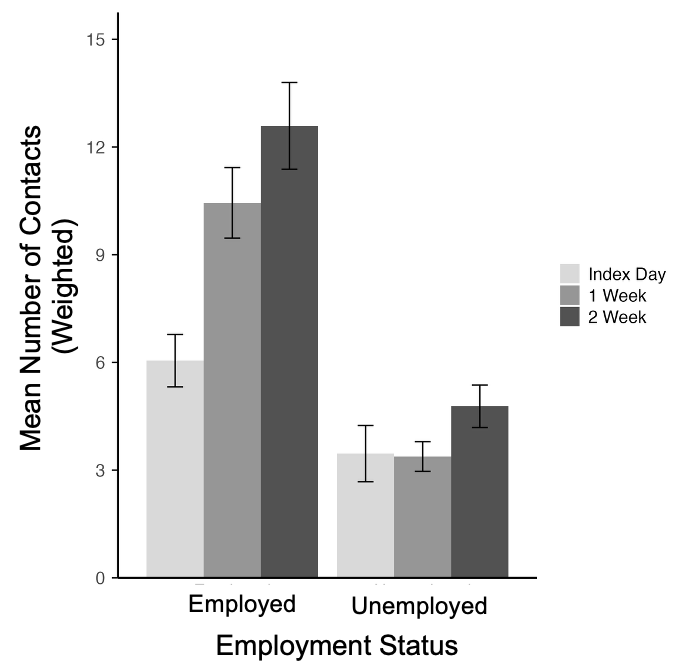

E. Household income (pre-tax)

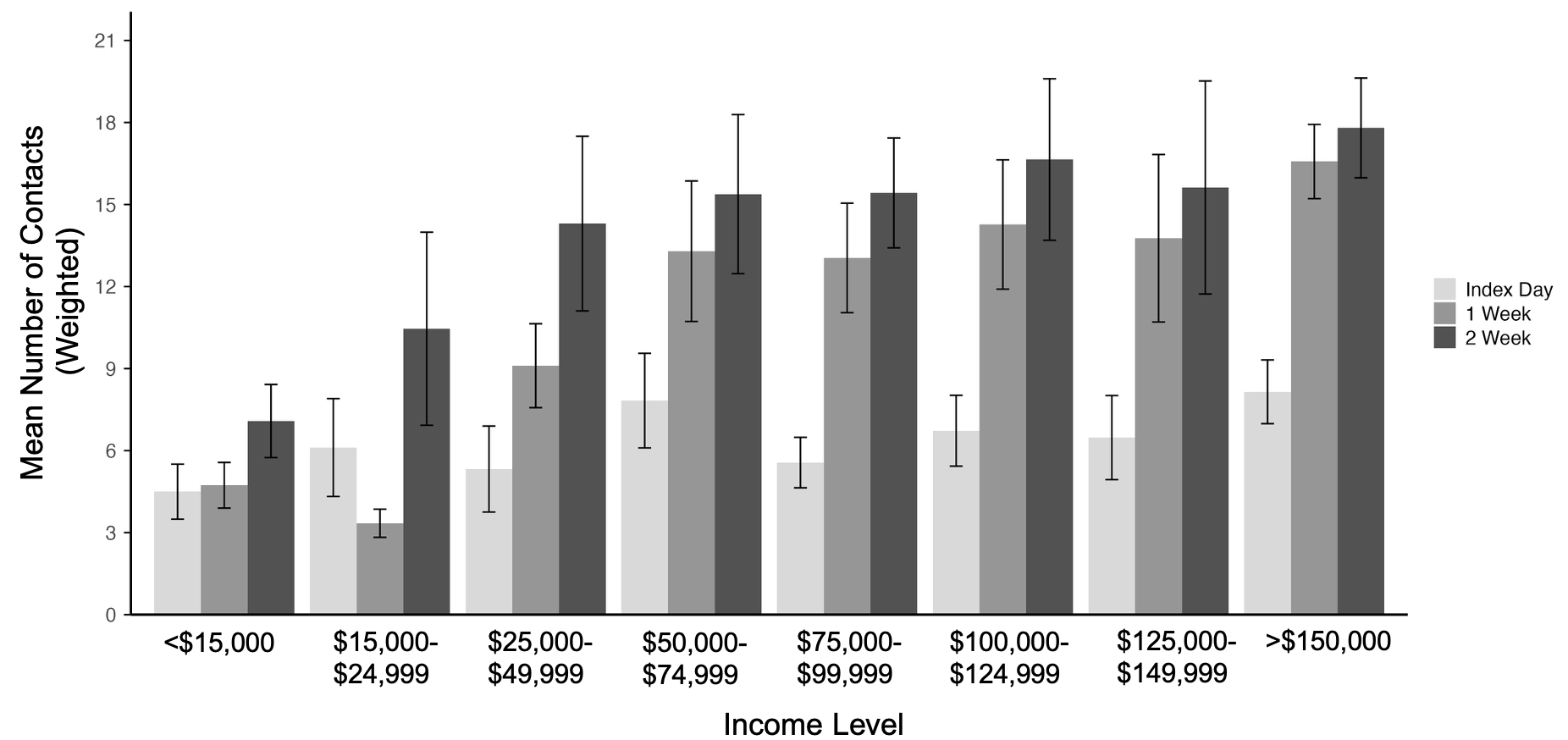

F. Occupation [N=508 (95% CI: 479–537)]	G. School attendance

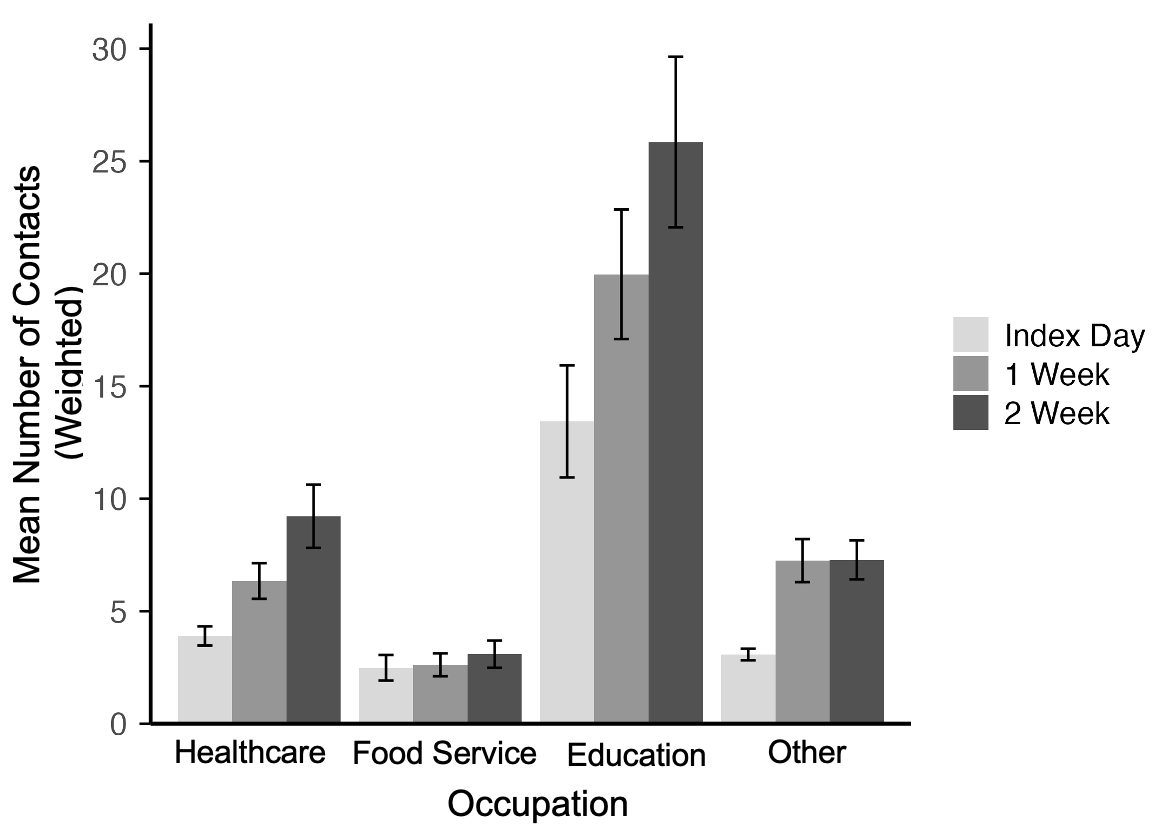

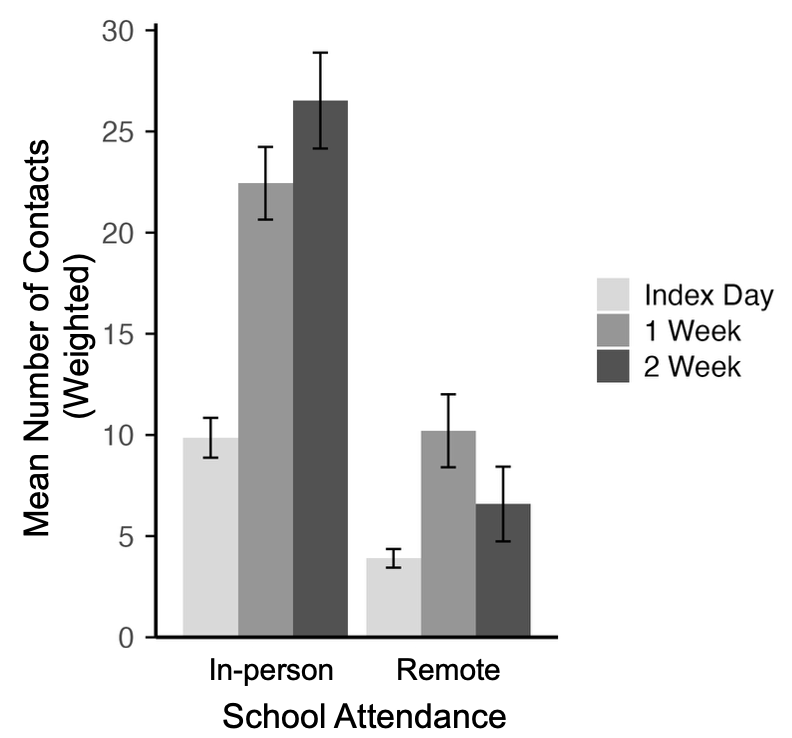

H. Residential setting	I. Healthcare encounter type

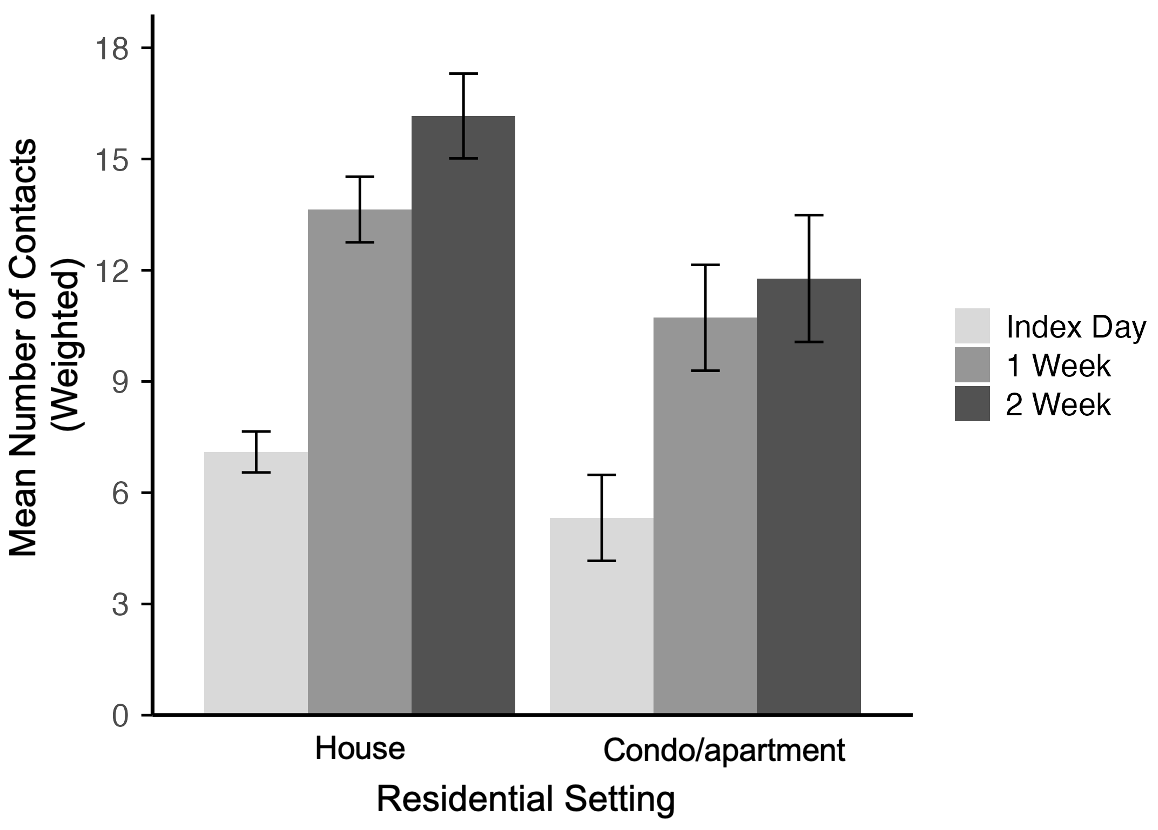

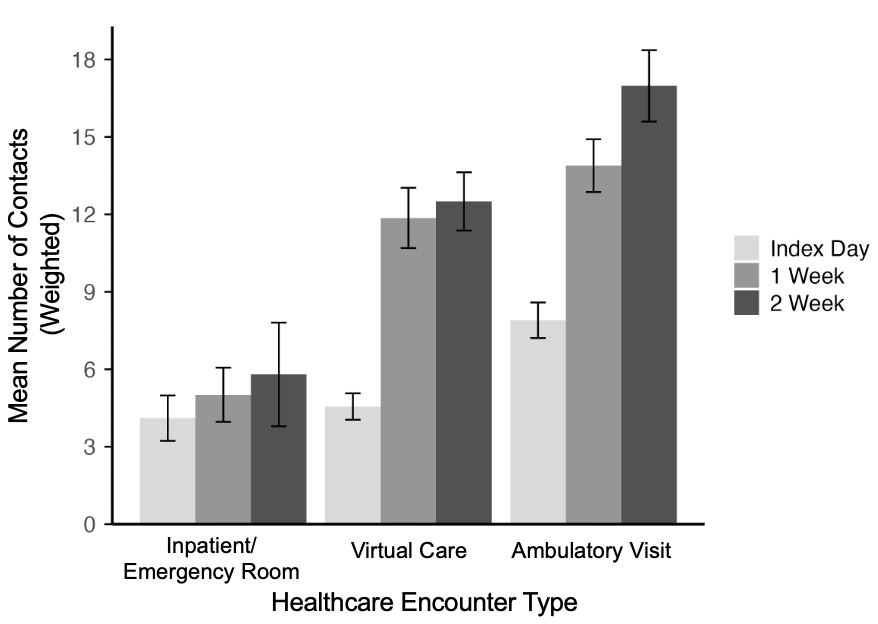

J. Disease type

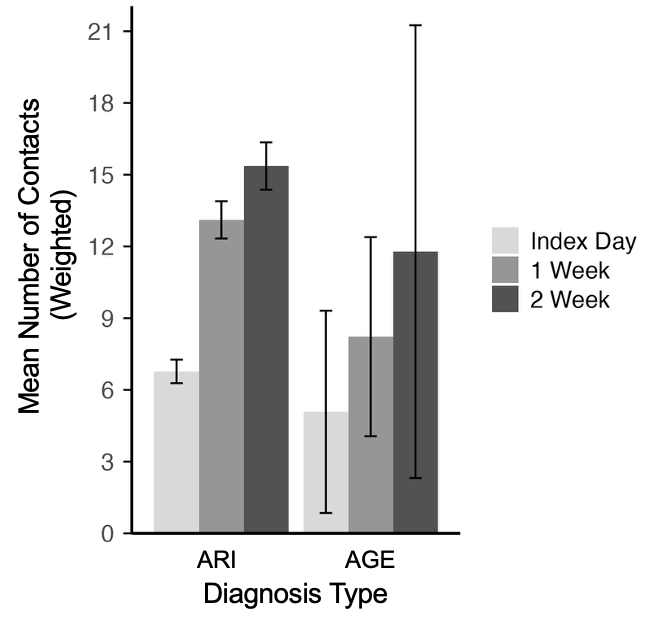

Abbreviations: AGE: Acute gastroenteritis, ARI: Acute respiratory infection.

Panels display weighted mean contacts for cases from 500 bootstrap resampled iterations of N=1,000. Vertical lines represent the standard deviation of the bootstrap means. Data from study participants that completed all longitudinal surveys were used in all panels.

*Other: American Indian or Alaska Native/Native Hawaiian or other Pacific Islander/Other

#### Supplementary Figure 3. Mean number of social contacts reported by acute gastroenteritis and acute respiratory infection cases over time by the timing of survey completion.

A. Unweighted

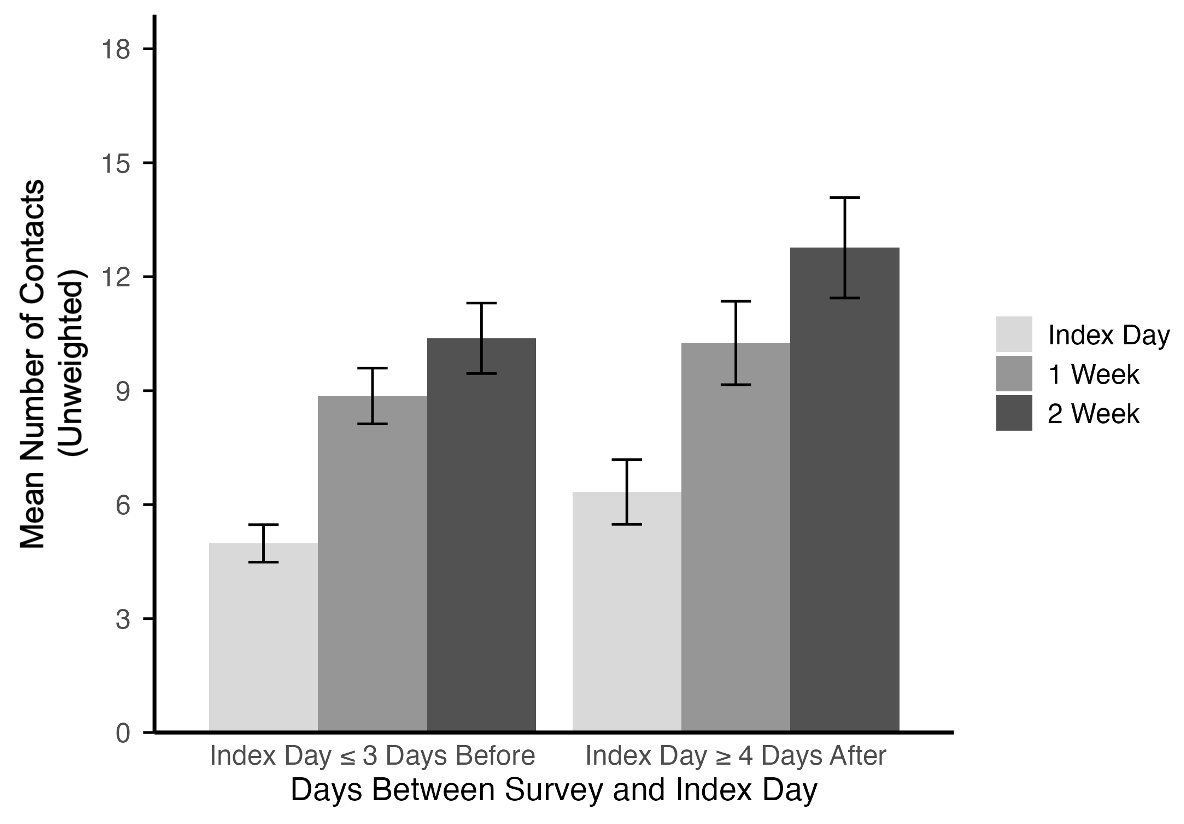

B. Weighted

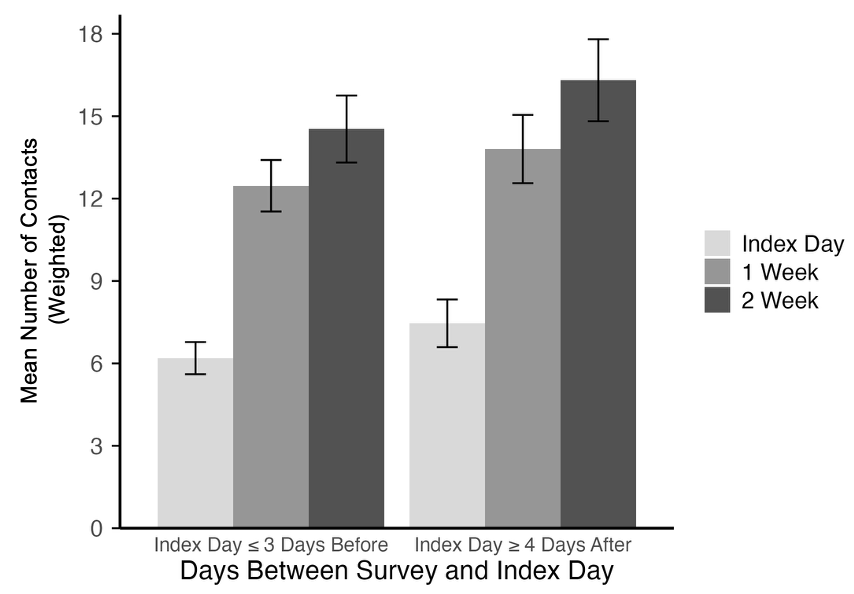

Panel A displays unweighted mean contacts for the observed analytic sample (N=1,000), with vertical lines representing error bars. Panel B displays weighted mean contacts for acute gastroenteritis and acute respiratory infection cases and their contacts from 500 bootstrap resampled iterations of N=1,000, with vertical lines representing the standard deviation of the bootstrap means.

#### Supplementary Figure 4. Weighted mean number of social contacts reported by acute gastroenteritis and acute respiratory infection cases over the course of infection across contact types and settings.

A. Contacts between cases and their household members at home

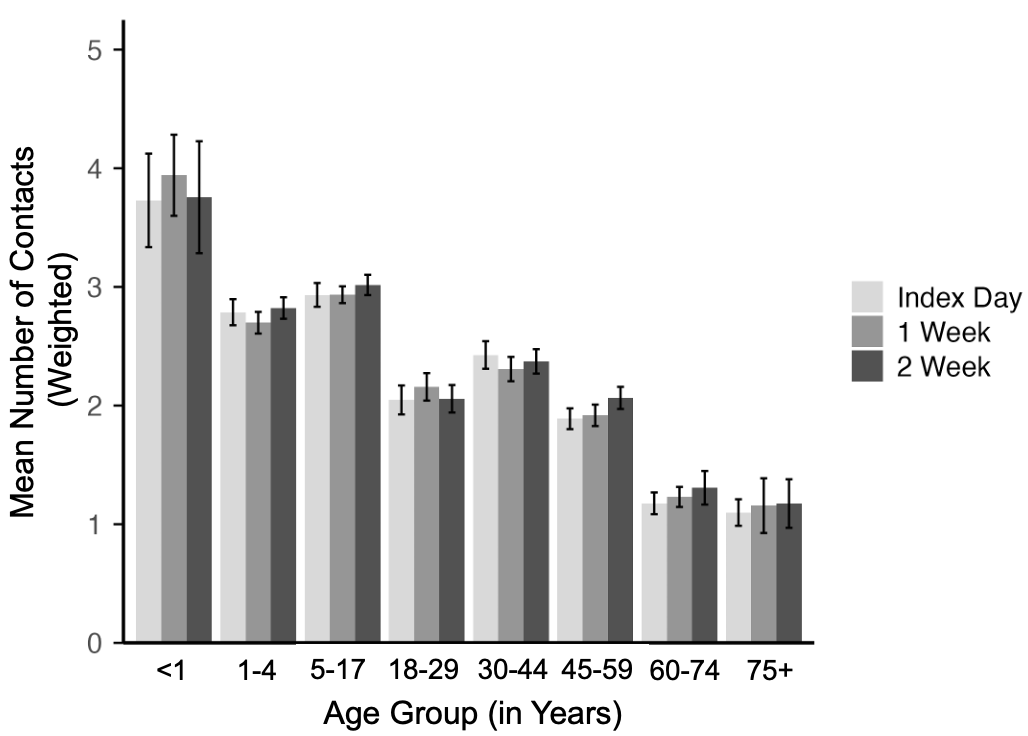

B. Contacts between cases and non-household members at home

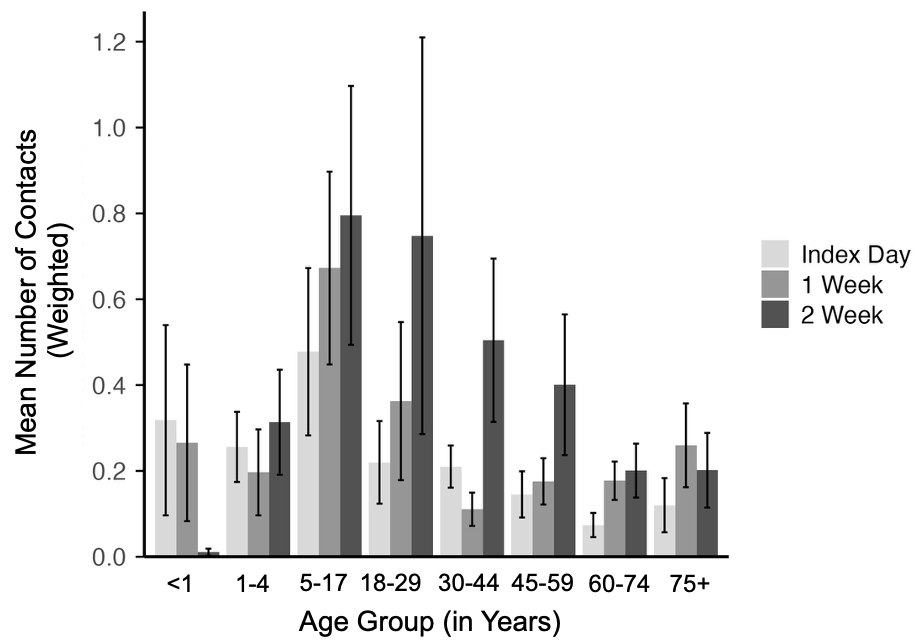

C. Contacts between cases and non-household members at work, school, and other settings

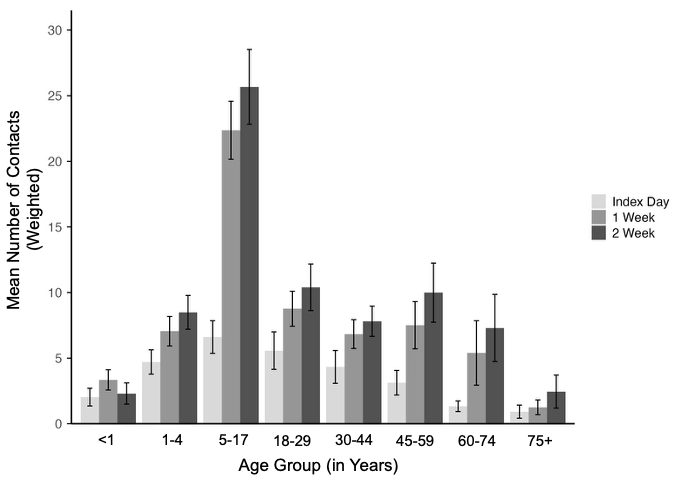

Panels display weighted mean contacts for cases from 500 bootstrap resampled iterations of N=1,000. Vertical lines represent the standard deviation of the bootstrap means. Data from study participants that completed all longitudinal surveys were used in all panels.

#### Supplementary Figure 5. Number of social contacts reported by household members over time by self-reported health status (N=691).

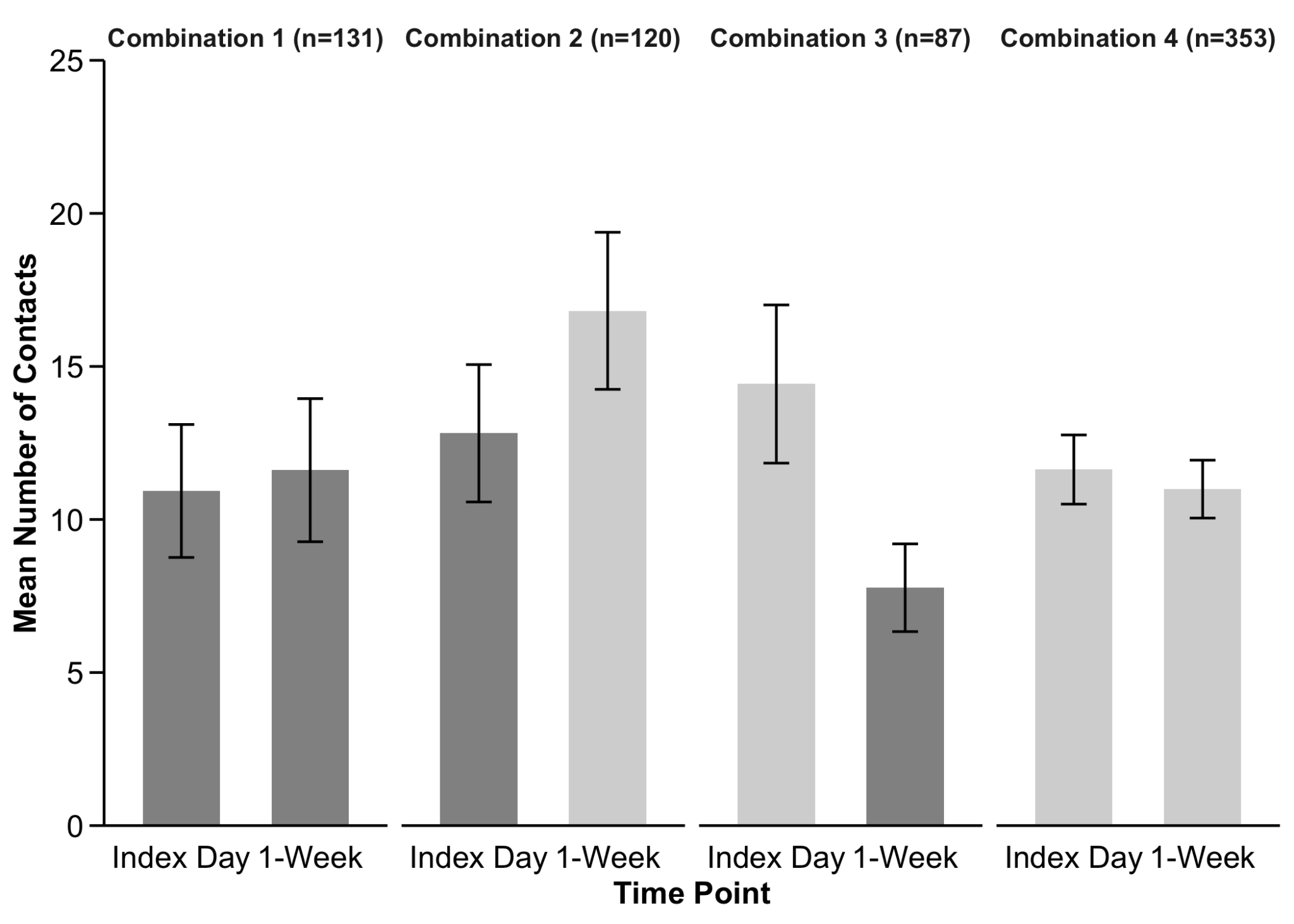

This figure presents the distribution of the number of contacts reported by household members of acute gastroenteritis/acute respiratory infection cases on the index date and at the 1-Week Follow-Up, stratified into the following four mutually exclusive health status combinations:

- Combination 1 includes household members who reported feeling sick on both days.
- Combination 2 includes those who felt sick on the index date but well at follow-up.
- Combination 3 includes those who felt well on the index date but sick at follow-up.
- Combination 4 represents individuals who reported feeling well on both days.

Each combination displays two bars—one for each time point—showing the mean number of contacts. Bars in darker grey represent data when household members felt sick, and those in lighter grey represent data when household members felt well. Vertical lines represent standard error bars. Only household members who provided data related to self-reported health status are included in this figure (N=691).

#### Supplementary Figure 6. Durations and average numbers of social contacts reported by household members over time by settings (N=709).

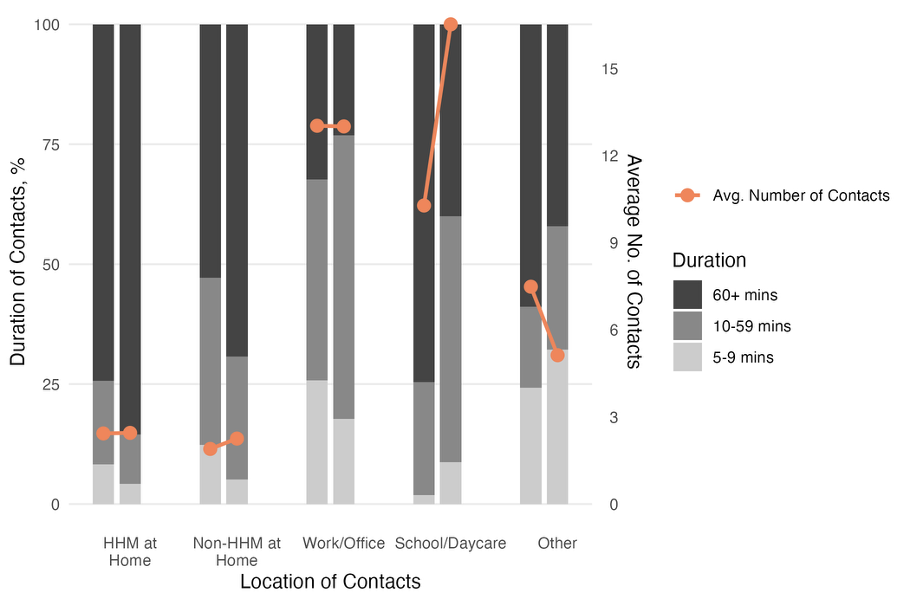

Abbreviation: HHM, household members.

Colored bars represent the proportion of social contact durations (5–9, 10–59, and 60+ minutes) (left axis). For each group of two bars, the left bar represents contacts made at index date and the right bar represents contacts made at one-week follow-up. The orange data points indicate the average number of social contacts in each setting over time (right axis).

#### Supplementary Figure 7. Weighted mean number of contacts reported by acute gastroenteritis and acute respiratory infection cases over the course of infection by infection mitigation behaviors.

A. Index survey

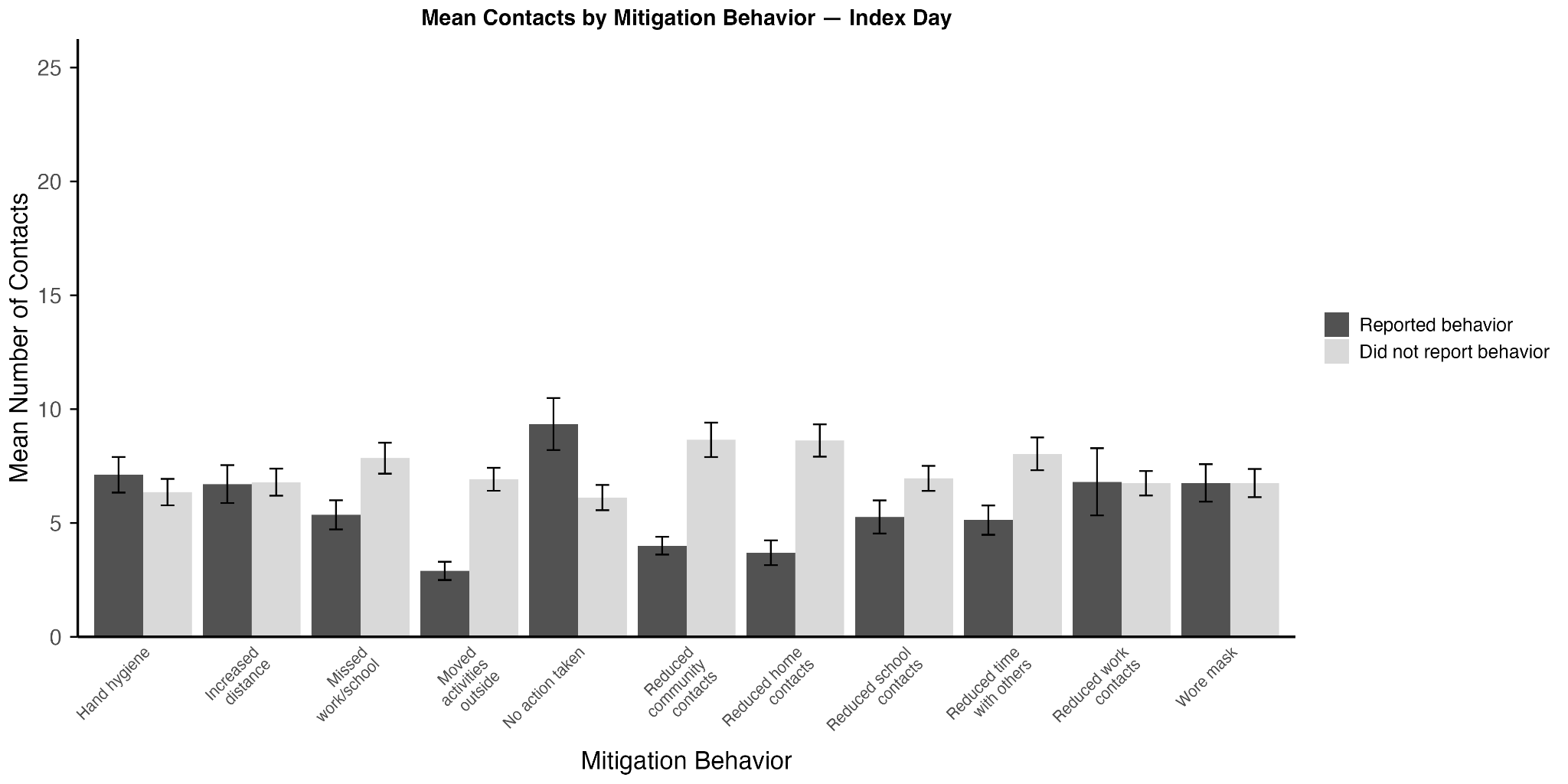

B. 1 week follow-up survey

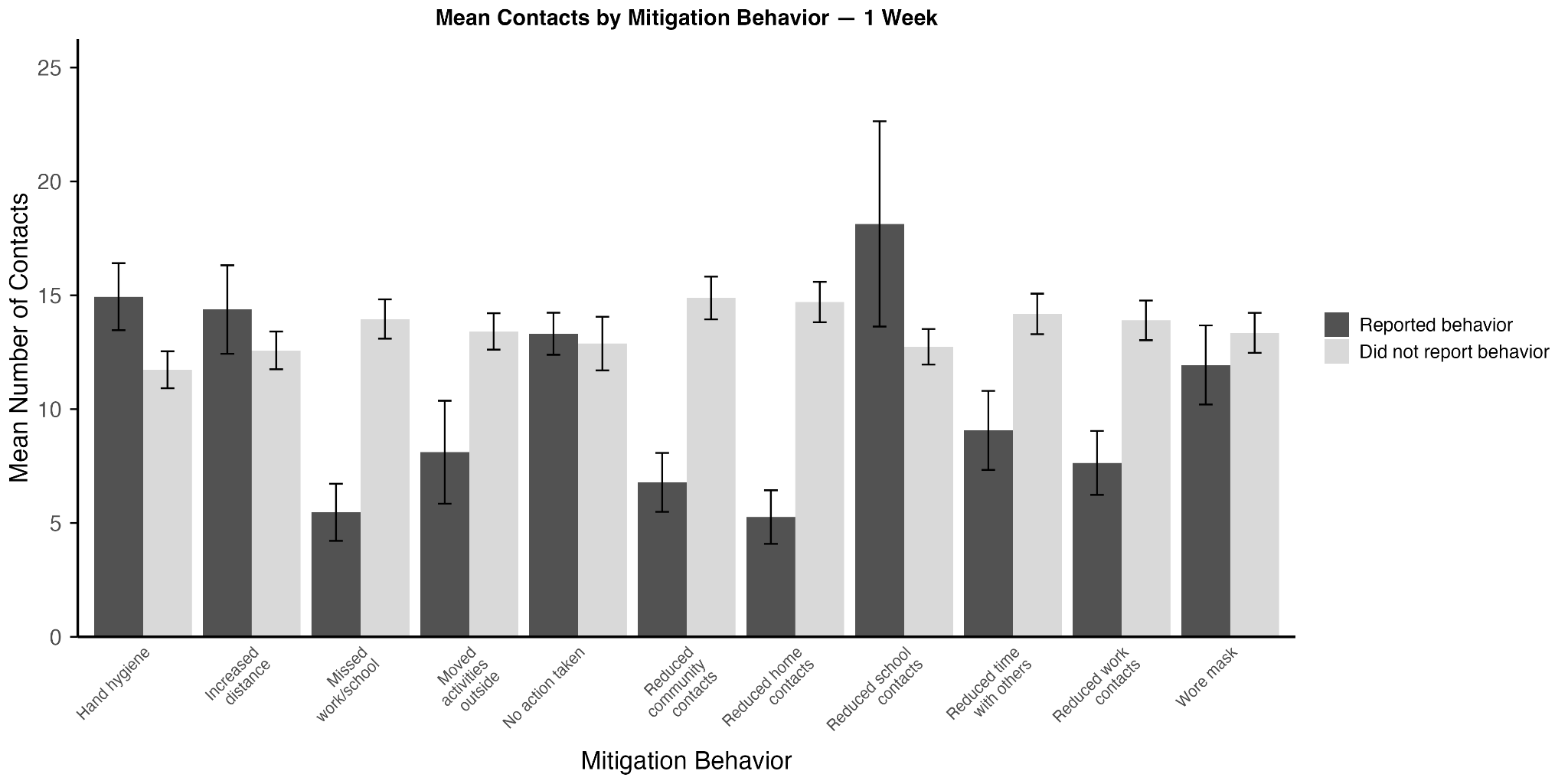

C. 2 week follow-up survey

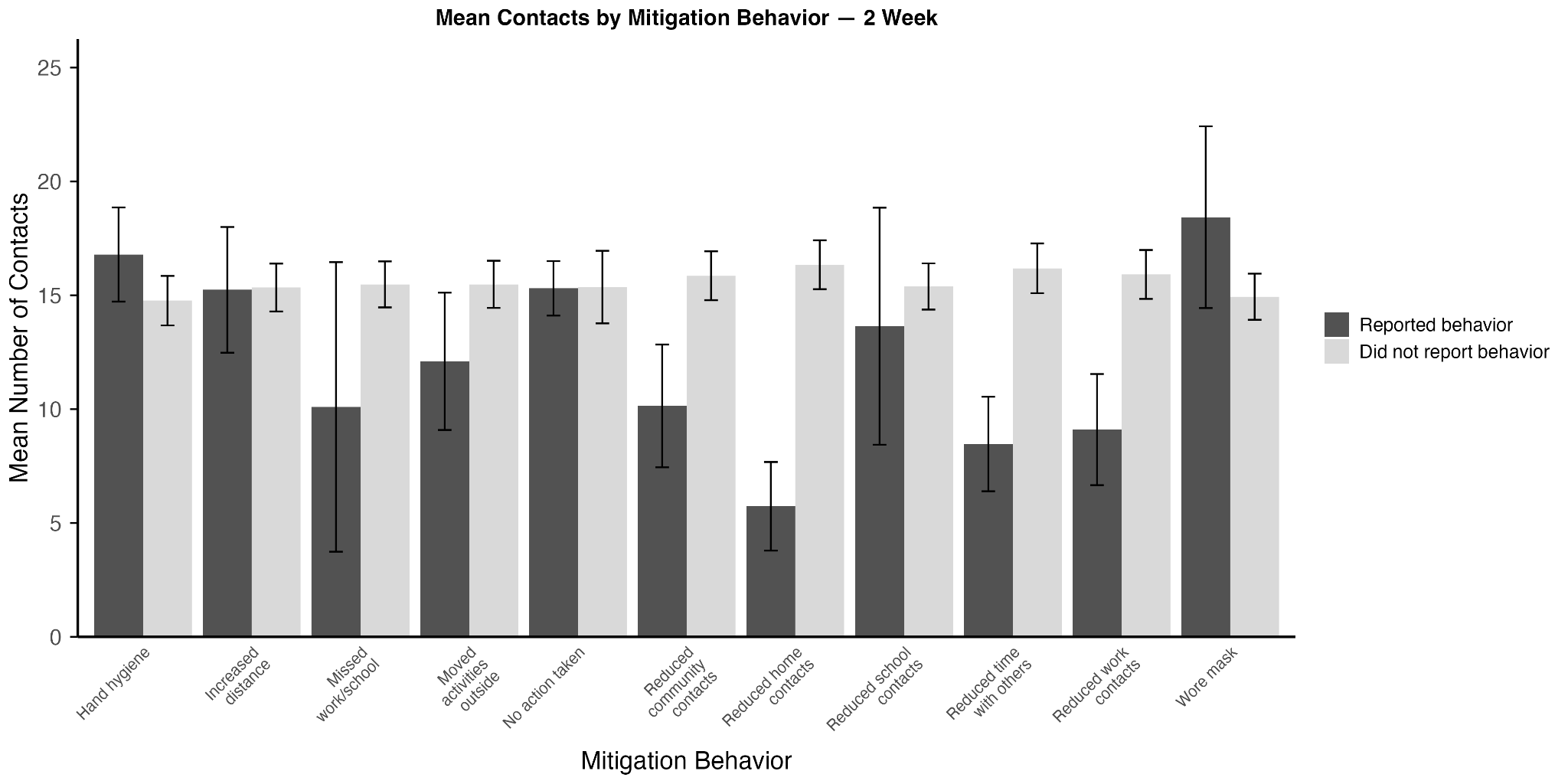

Each plot displays weighted mean contacts for cases from 500 bootstrap resampled iterations of N=1,000. Vertical lines represent the standard deviation of the bootstrap means.

#### Supplementary Figure 8. Unweighted mean number of social contacts reported by acute gastroenteritis and acute respiratory infection cases over the course of acute infection by characteristics and settings.

A. Mean contacts by survey	B. Mean contacts by age

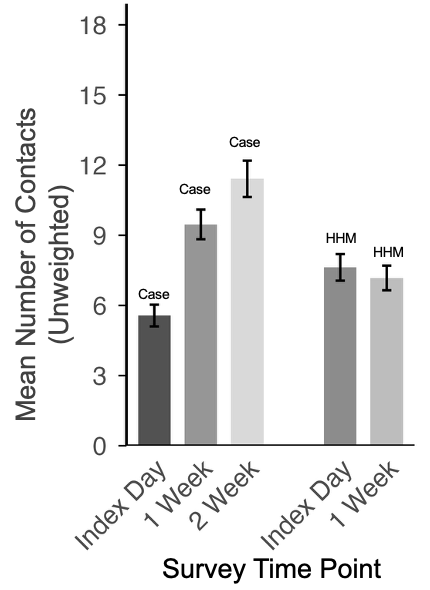

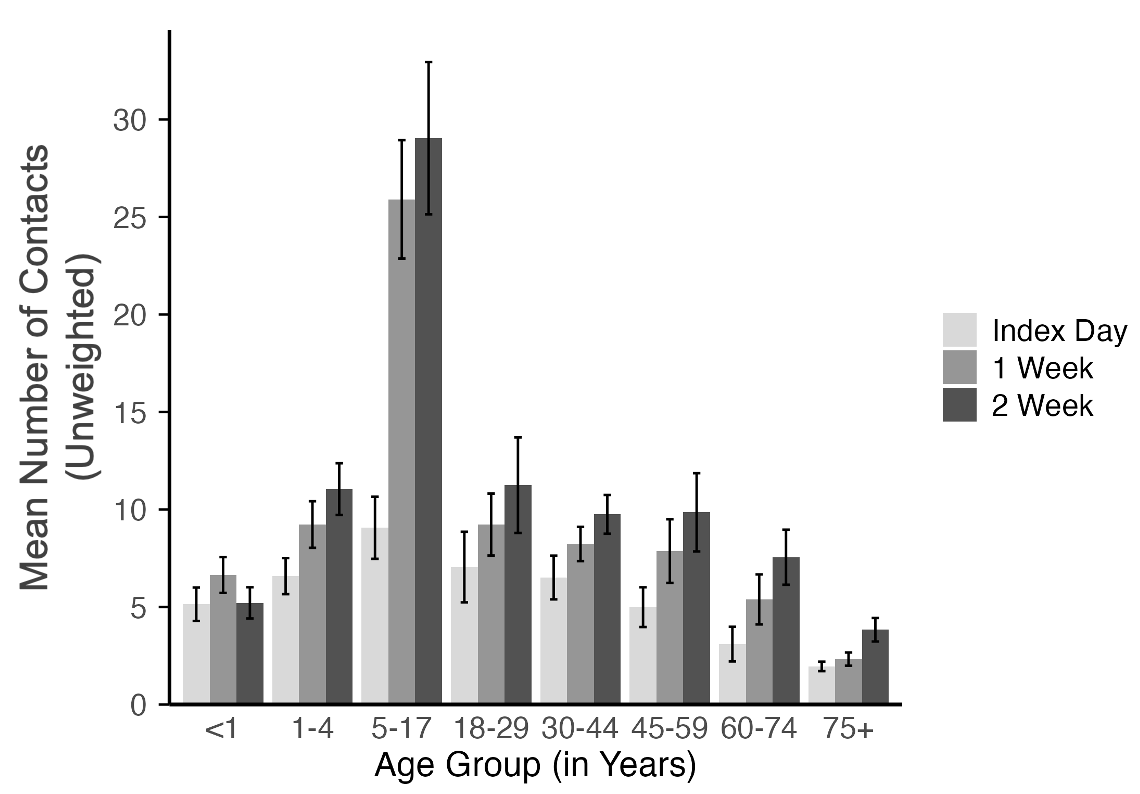

C. Unweighted social contact matrices (mean number of contacts) between acute gastroenteritis and acute respiratory infection cases and their contacts over time by age group (N=1,000)

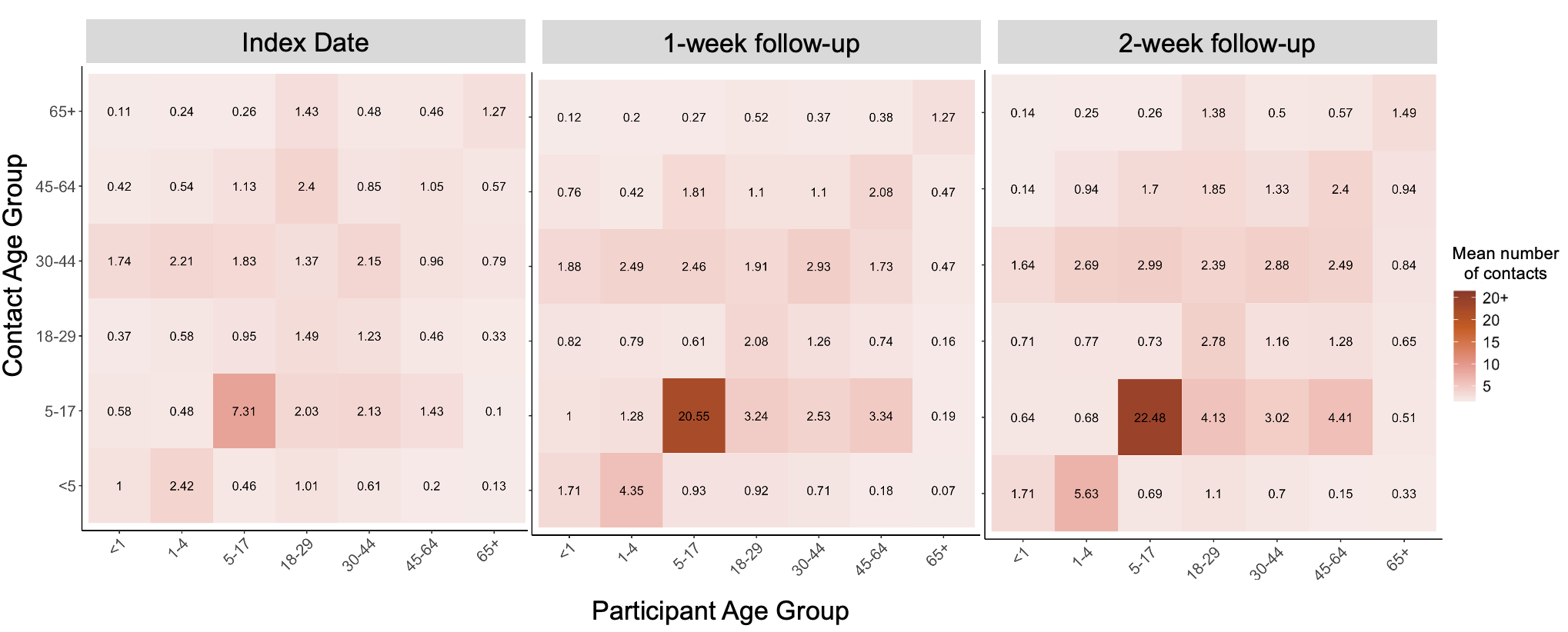

D. Durations and average numbers of social contacts over the course of illness by setting^1^

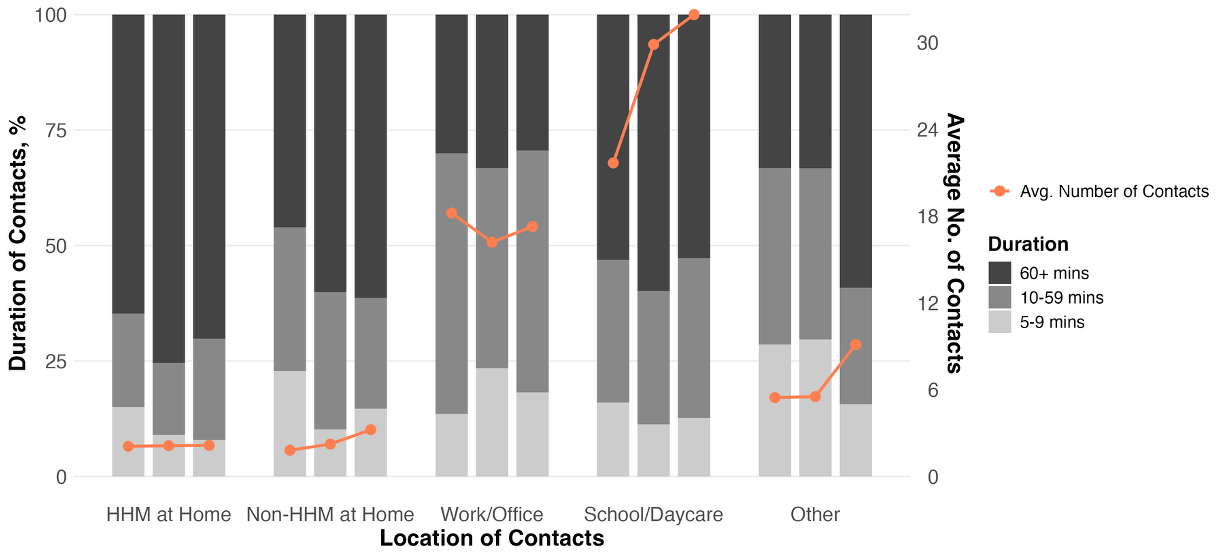

E. Number of social contacts reported by cases over time by self-reported health status (N=994)^2^

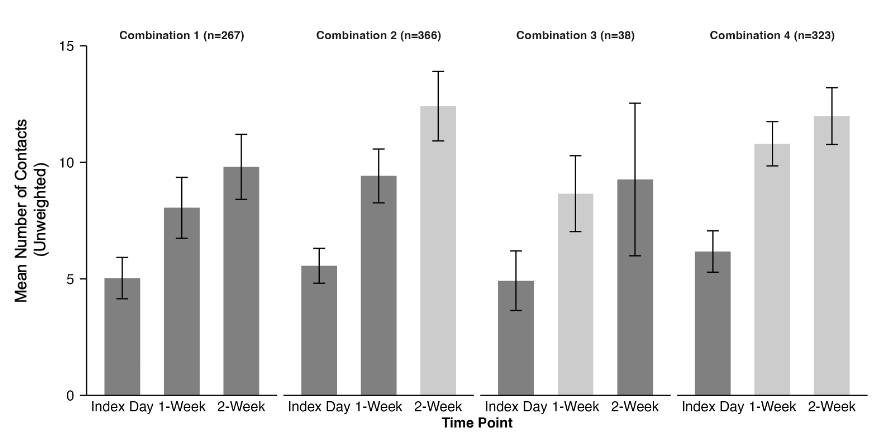

F. Mean contacts by sex	G. Mean contacts by racial group

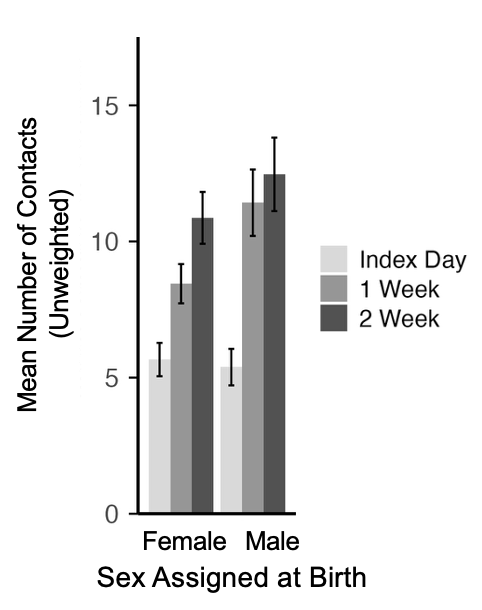

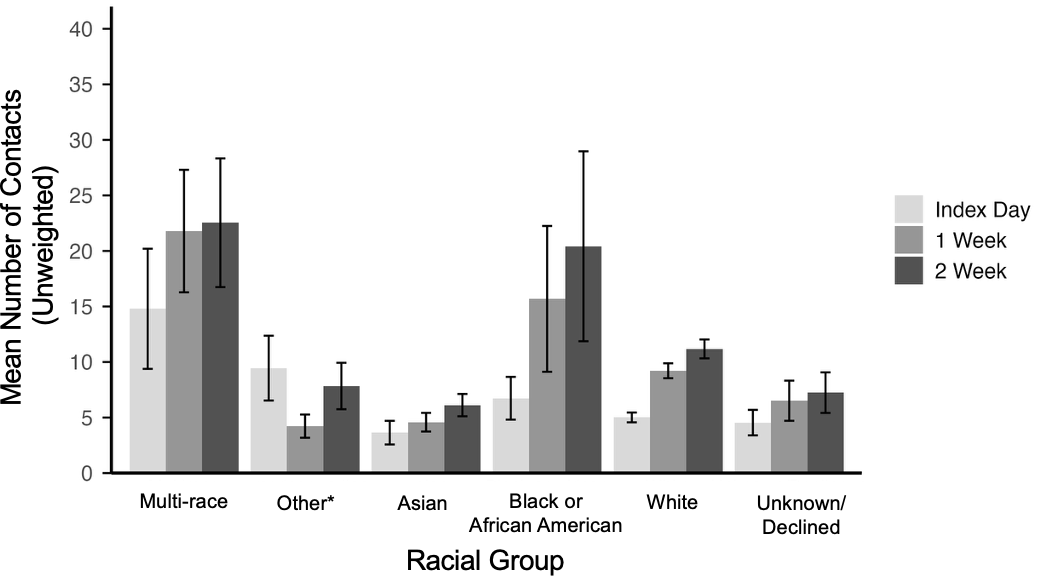

H. Mean contacts by ethnicity	I. Mean contacts by employment status

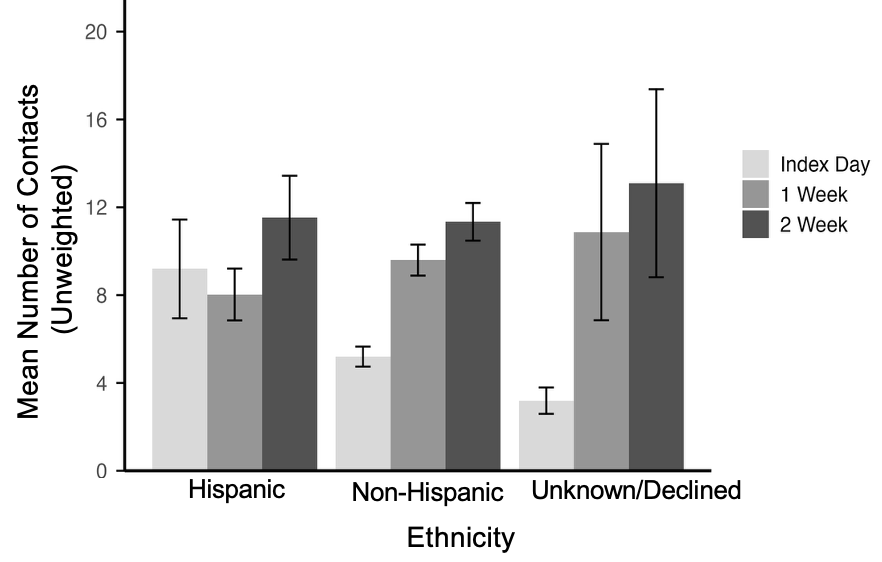

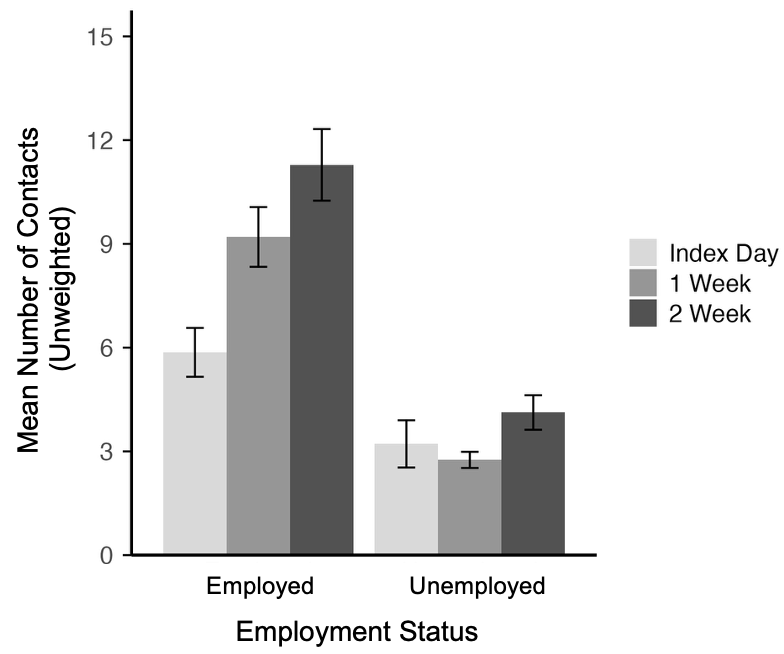

J. Mean contacts by income level

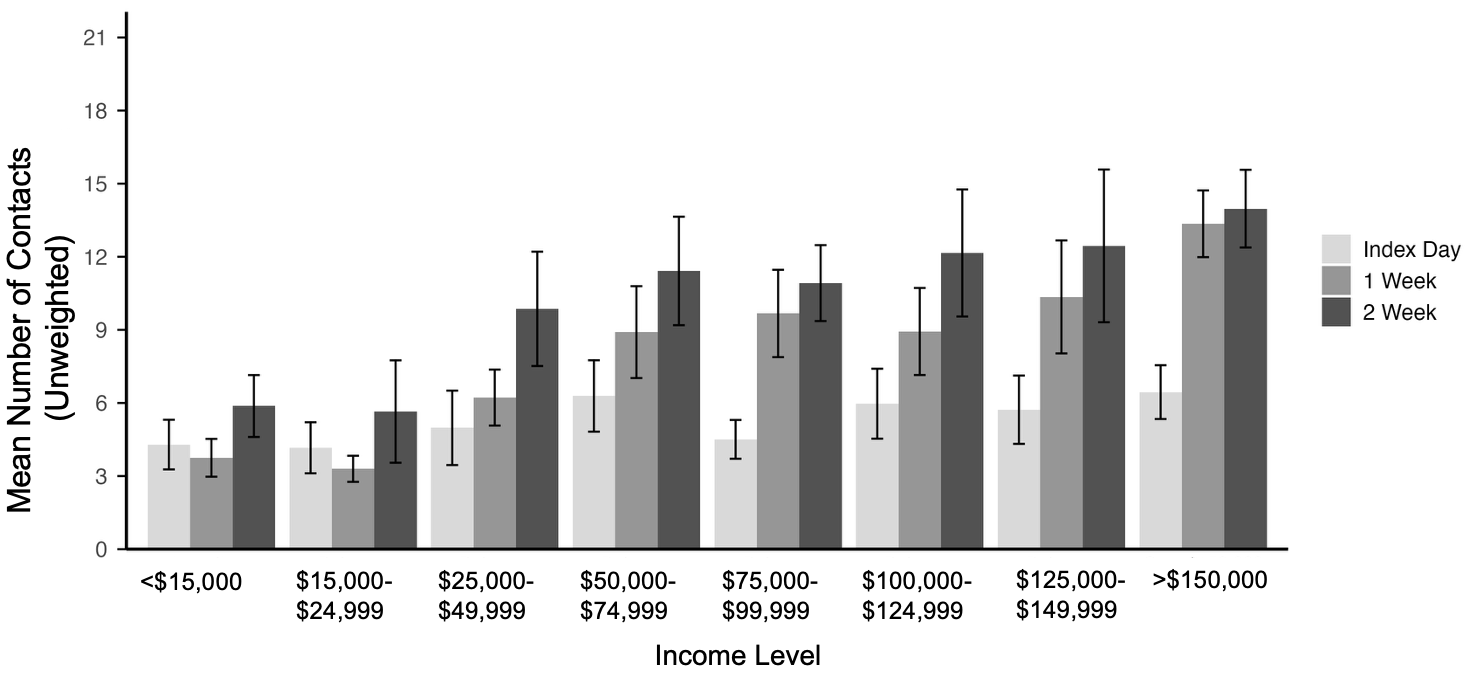

K. Mean contacts by occupation	L. Mean contacts by school attendance

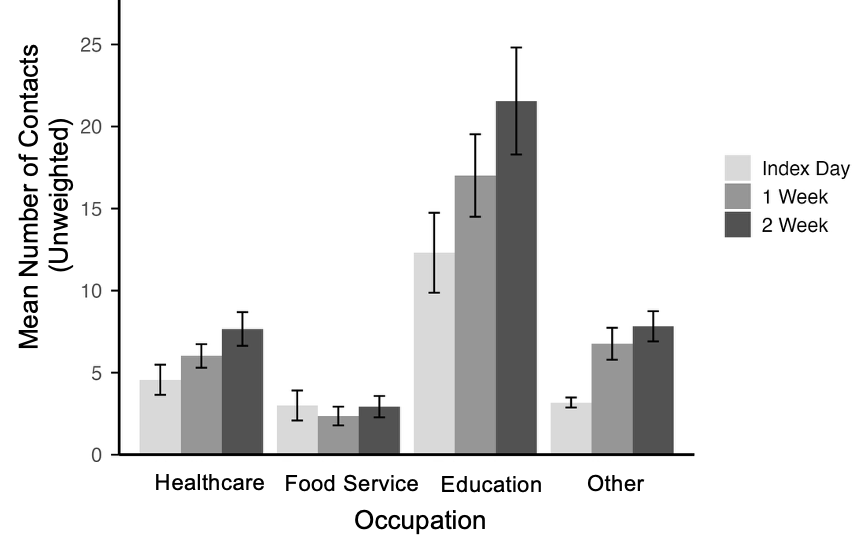

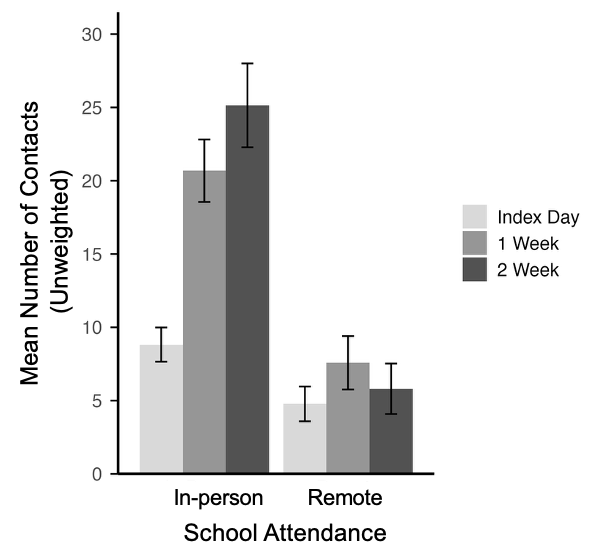

M. Mean contacts by home setting	N. Mean contacts by healthcare type

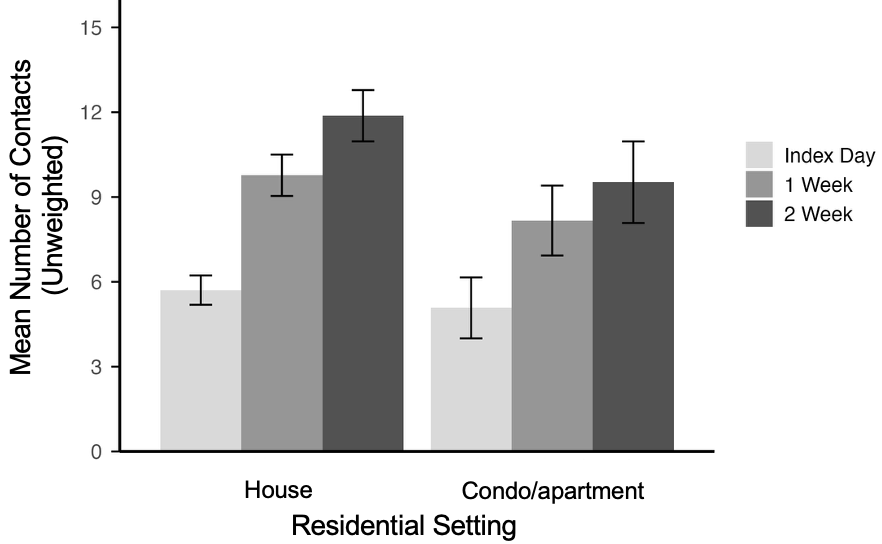

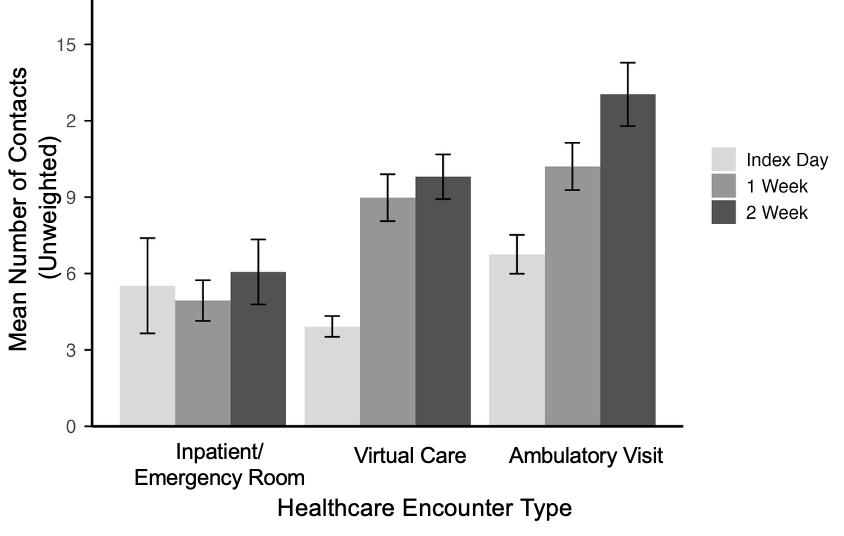

O. Mean contacts by diagnosis type

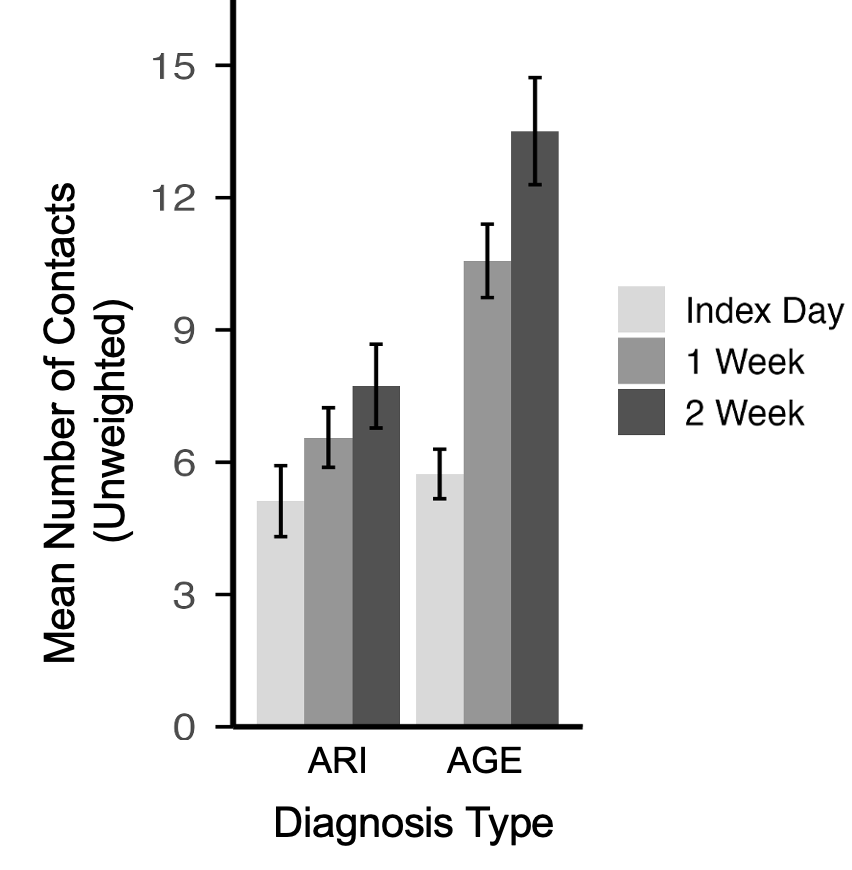

P. Mean contacts between acute gastroenteritis and acute respiratory infection cases and their household members at home

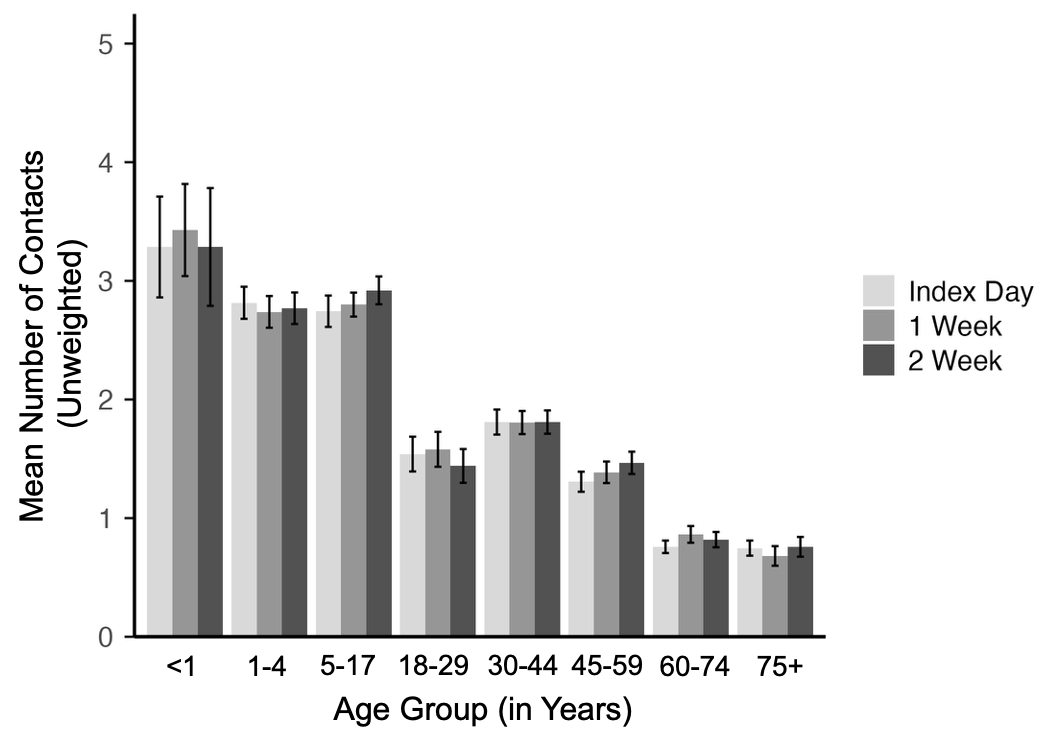

Q. Mean contacts between acute gastroenteritis and acute respiratory infection cases and non-household members at home

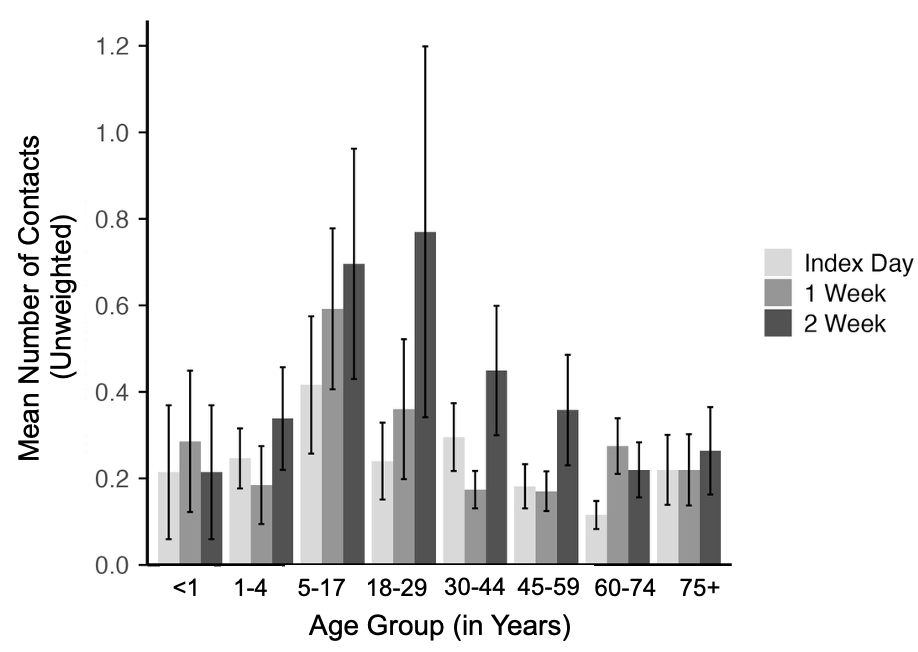

R. Mean contacts between acute gastroenteritis and acute respiratory infection cases and non-household members at school, work, and other settings

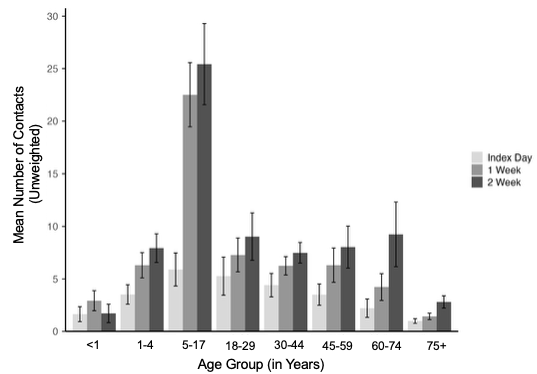

Abbreviations: HHM: household members; AGE: Acute gastroenteritis; ARI: Acute respiratory infection.

Vertical lines represent standard error bars.

^1^The figure displays unweighted mean contacts for acute gastroenteritis and acute respiratory infection cases (N=1,000). Colored bars represent the proportion of social contact durations (5-9, 10-59, and 60+ minutes) (left axis). For each group of three bars, the left bar represents contacts made at index date, the middle bar represents contacts made at one-week follow-up, and the right bar represents contacts made at two-week follow-up. The orange data points indicate the average number of social contacts in each setting over time, with vertical lines representing standard error bars (right axis).

^2^This figure presents the distribution of the number of contacts reported by acute gastroenteritis and acute respiratory infection cases on the index date, 1-Week-, and 2-Week Follow-Up, stratified into the following four mutually exclusive health status combinations:

- Combination 1 includes cases who reported feeling sick on all 3 days.
- Combination 2 includes those who felt sick on the index date and 1-Week Follow-Up but well at the 2-Week Follow-Up.
- Combination 3 includes those who felt sick on the index date, well at the 1-Week Follow-Up, and sick again at the 2-Week Follow-Up.
- Combination 4 includes those who were sick on the index date, but reported feeling well at the 1- and 2-Week Follow-Up.

Each combination displays three bars–one for each time point–showing the mean contacts and vertical lines representing standard error bars. Bars in darker grey represent data when cases felt sick, and those in lighter grey represent data when cases felt well. Six participants who completed all 3 surveys were excluded from this figure due to missing data related to self-reported health status.

### Supplementary Methods

*AGE and ARI case recruitment and eligibility criteria*

Eligible individuals were invited to complete an online consent form and an online enrollment survey, which determined their eligibility. Cases were excluded from the study if their symptoms were chronic (lasting more than three weeks) or if they resided in long-term care facilities, including hospices, nursing homes, skilled nursing facilities, or assisted living facilities. If symptom onset was ≥14 days prior to the enrollment survey date, they were excluded. If the index date was ≥6 days prior to the enrollment survey date, participants were ineligible to proceed to the second part of the survey due to concerns about recall bias.

Each participant was only allowed to enroll in the study once. Individuals who participated as an AGE or ARI case were not eligible to participate again as a household member, even if other members of their household also became ill. Likewise, participants enrolled as AGE cases were not eligible to later participate as ARI cases, and vice versa.

Recruitment of AGE and ARI cases occurred from March 18 to June 30, 2024, and from October 14, 2024, to March 31, 2025, and data collection continued through May 31, 2025, to ensure that all enrolled participants had sufficient time to complete the longitudinal surveys.

*Definition of social contacts*

We defined social contacts as interactions lasting more than five minutes for several reasons. Prior social contact studies suggested that contact duration was related to the intensity and nature of social interactions. For example, Mossong, *et al.* found that contacts lasting less than five minutes were predominantly non-physical, whereas longer contacts were more likely to involve physical contact and to occur repeatedly (*7*). Contact duration may also be epidemiologically relevant, as transmission risk generally increases with the duration of exposure, as demonstrated in empirical contact-network modeling studies such as Toth, *et al.* The previous study also found that brief contacts were less reliably recalled and reported in contact surveys (*29*). Smieszek, *et al.* compared self-reported contacts with sensor-measured proximity and found lower reporting of contacts lasting less than five minutes than of longer contacts (*31*). A five-minute threshold has been used in infectious disease transmission research; for example, Potter et al. defined social contacts for influenza transmission modeling as interactions involving close proximity for approximately five minutes or longer (*30*). For these reasons, in our study, we sought to minimize response burden by asking them to report substantive social interactions rather than attempting to enumerate every brief or incidental encounter, especially because participants were acutely ill at the time of the index survey.

*Household member recruitment*

To recruit household members to the SickMix study, AGE and ARI cases were asked to provide information on up to five household members in the enrollment survey. If a case had more than five household members, they were asked to select up to five household members to invite.
